# Genetic associations with EEG signatures in mild cognitive impairment: insights into Alzheimer’s disease pathophysiology

**DOI:** 10.64898/2026.08.25.26361294

**Authors:** Ari M. Siitonen, Timo Saarinen, Mia Liljeström, Antti S. Kinnunen, Verna Heikkinen, Mahshid Pashootan, Niina Kanerva, Shrikanth Kulashekhar, Ville Mäntynen, Jaakko Hotta, Anne Koivisto, Irina Anurova, Maria J. Lagartos-Donate, Fernando Maestú, Camillo Marra, Paolo M. Rossini, Ira A. Haraldsen, Hanna Renvall

**Affiliations:** BioMag Laboratory, HUS Diagnostic Center, Helsinki University Hospital, University of Helsinki, and Aalto University School of Science, Helsinki, Finland; HUS Diagnostic Center, Clinical Neurophysiology, Clinical Neurosciences, Helsinki University Hospital and University of Helsinki, Helsinki, Finland; Department of Neuroscience and Biomedical Engineering, Aalto University, Finland; Bittium Biosignals, Kuopio, Finland; Department of Neurology, Helsinki University Hospital and Clinical Neurosciences, Neurology, University of Helsinki, Helsinki, Finland; Department of Geriatrics, Neurological Outpatient Polyclinic and BioMag Laboratory, Helsinki University Hospital, Helsinki, Finland; Department of Neurosciences, Faculty of Medicine, University of Helsinki, Helsinki, Finland; NeuroCenter, Kuopio University Hospital, Kuopio, Finland; Pre Diagnostics AS, Oslo, Norway; Laboratory of Molecular Neuroscience, Division of Anatomy, Department of Molecular Medicine, Institute of Basic Medical Sciences, University of Oslo, Oslo 0372, Norway; Centre for Cognitive and Computational Neuroscience, Universidad Complutense de Madrid, Madrid, Spain; Department of Experimental Psychology, Cognitive Psychology and Speech and Language Therapy, Universidad Complutense de Madrid, Pozuelo de Alarcón, Spain; Institute of Sanitary Investigation (IdISSC), San Carlos University Hospital, Madrid, Spain; Memory Clinic, Fondazione Policlinico Universitario Agostino Gemelli IRCCS, Rome, Italy; Department of Neuroscience, Catholic University of the Sacred Heart, Rome, Italy; Department of Neuroscience and Neurorehabilitation, IRCCS San Raffaele, Rome, Italy; Department of Neurology, Oslo University Hospital, Oslo, Norway

**Author notes:** Correspondence to: Ari Siitonen, BioMag Laboratory, HUS Diagnostic Center, Helsinki University Hospital FI-00029 HUS, FINLAND.

**Keywords:** mild cognitive impairment continuum, Alzheimer’s disease, electroencephalography, genetics, Bayesian reduced rank regression

## Abstract

The mild cognitive impairment (MCI) continuum represents a critical stage in the Alzheimer’s disease (AD), yet much of the genetic determinants underlying early neural dysfunction in AD remain unclear. Electroencephalography (EEG)-derived features as heritable markers of neuronal network activity may provide biologically informative endophenotypes for studying cognitive decline in the MCI.

We investigated associations between functional genetic variation and resting-state EEG features in 169 Finnish individuals with symptoms spanning from subjective cognitive decline to MCI. Periodic alpha- and beta-band features and aperiodic spectral parameters were derived from eyes-closed EEG recordings. Genome-wide genotypes were quality-controlled, imputed, functionally annotated (yielding 16,935 gene regions comprising 90,607 functional variants), and associated with latent EEG features using Bayesian reduced rank regression. We integrated the results with neurobiology-related literature review and known AD-associated loci and performed unsupervised clustering of participants based on EEG-associated genetic variation, testing clinical and biomarker differences between clusters.

We identified 145 genes associated with periodic EEG features and 39 genes with aperiodic EEG features at *P* < 0.005, although no associations survived correction for multiple testing. Candidate genes converged on pathways for synaptic transmission, neuronal excitability and neurodevelopment (e.g., RASGEF1C and TREML2 mapped to loci previously associated with AD). The aperiodic-feature candidates were more enriched for neuroinflammatory processes (e.g., C5AR2 within a FinnGen AD-associated region) relative to the periodic-feature candidates which were more frequently involved in intracellular neuronal maintenance and signaling. Unsupervised clustering based on periodic EEG-associated variants delineated participant subgroups differing significantly in plasma p-tau217 concentrations and delayed verbal recall after multiple-testing correction. The top genes contributing to cluster formation included ACAN, INPP5B, CAMKK2, CABIN1 and SPATA13.

These findings suggest that periodic and aperiodic EEG features capture partly distinct biological processes within the MCI continuum. The convergence of candidate genes on synaptic, neurodevelopmental and neuroinflammatory pathways, and genetically informed clustering linked to plasma p-tau217 and delayed verbal recall, supports the use of EEG-derived endophenotypes for dissecting heterogeneity in early cognitive decline and AD-related pathology.

## Introduction

Mild cognitive impairment (MCI), a condition characterized by objectively tested cognitive decline but preserved daily functioning, affects an increasing share of the aging population, with a global prevalence rate of 20%.^1^Individuals with MCI are at heightened risk of progressing to dementia— particularly Alzheimer’s disease (AD)—with conversion rates ranging from 10% to 34%.^2^ Key deficits in the prodromal phase of AD typically emerge in the memory domain,^3-6^ although impairments in other cognitive domains such as executive function or language may also occur ^7^ and become increasingly pronounced as the disease progresses.^8-10^ Even subjective cognitive decline (SCD), a pre-MCI stage characterized by cognitive concerns without clear objective cognitive findings, is associated with an elevated risk of developing dementia,^11,12^ consequently shifting research focus towards this earlier at-risk group.^13,14^ The MCI continuum, starting from the subjective stage before fully developed MCI, thus provides a critical window for studying the genetic and other biological risks of AD. Yet current clinical methods are insufficient in identifying which individuals with SCD and MCI are most likely to progress to AD and clinical dementia, limiting targeted preventive care and pharmacological interventions.

AD pathophysiology unfolds years before overt changes in behavior or brain structures occur.^15,16^ The hallmarks of AD pathophysiology, accumulating amyloid-β plaques and tau-tangles in the brain,^17^ are reflected earlier in cerebrospinal fluid and plasma biomarkers, such as phosphorylated tau at threonine 217 (p-tau217).^18,19^ At the genetic level, AD risk is most prominently influenced by the APOEε4 allele.^20^ While 75 genome-wide risk loci have been associated with AD, and polygenic risk score shows a modest but significant association with progression from MCI to AD,^21^ a substantial gap exists between twin-study based heritability estimates of AD (70%) and genome-wide association studies (GWAS) (3.1%). This discrepancy implies that many causal variants affecting the disease’s pathophysiology remain unidentified.^22^ Bridging this gap may require intermediate phenotypes, such as specific neurophysiological signatures, which could provide a more direct readout of the early neural dysfunction than clinical outcomes can provide.

Synaptic dysfunction is considered one of the earliest pathological changes in AD.^23^ Electroencephalography (EEG) and magnetoencephalography (MEG) directly capture neuronal population activity with high temporal resolution, providing a non-invasive means of detecting early network-level disturbances. Indeed, alterations in electrophysiological activity are detectable already during the preclinical and prodromal stages of AD and correlate with cognitive decline.^24,25^ Among the most consistent findings are slowing and reduced power of the dominant posterior alpha (7–13 Hz) rhythm, which have been shown to reflect disrupted large-scale cortical synchronization.^26,27^ Furthermore, alterations in beta-band (13–30 Hz) activity and aperiodic spectral characteristics may capture complementary aspects of AD-related network dysfunction. Beta-band activity has been associated with cortical inhibitory dynamics and GABAergic processing,^28^ and changes in beta-band activity have been reported along the AD continuum.^29^ In contrast, aperiodic spectral features, particularly the 1/f spectral exponent, are thought to reflect excitation–inhibition (E-I) balance and arousal-related neural activity,^30,31^ and correlate with amyloid-β and tau pathology.^32^ Importantly, these spectral EEG/MEG measures are relatively stable across time and show substantial heritability,^33-36^ supporting their utility as genetically informative endophenotypes.

Here, we investigated how genetic variation modulates EEG signatures in a Finnish cohort spanning the MCI continuum and evaluated whether these electrophysiological metrics can serve as endophenotypes for cognitive performance, thereby helping to identify genetic risk factors for cognitive decline. Specifically, we examined whether associations between EEG features and cognitive profiles manifest at the genetic level, by evaluating the degree to which these candidate genes overlap with established AD pathophysiology. We focused on the periodic oscillatory activity in the 7–13 Hz (alpha) and 13–30 Hz (beta) frequency ranges, alongside aperiodic EEG features, to explore how common genetic variation influences these prominent electrophysiological measures and contributes to domain-specific cognitive performance in individuals with MCI.

## Material and methods

### Study subjects

We enrolled 187 participants, aged between 60 and 80 years (mean age 68 ± 5 years; 58 % females), from the Finnish cohort participating in the AI-Mind project (Haraldsen et al., 2024), spanning a cognitive continuum from subjective cognitive decline (42% of the participants, N=78) to objective mild cognitive impairment (58%, N=109). The descriptive statistics are shown in **Table 1**. The participants were mainly (89%) recruited via the Helsinki University Hospital (HUS) neurology outpatient clinics; the rest of the participants were recruited through community outreach with prescreening of suitability and inclusion to the study.

**Table 1.**
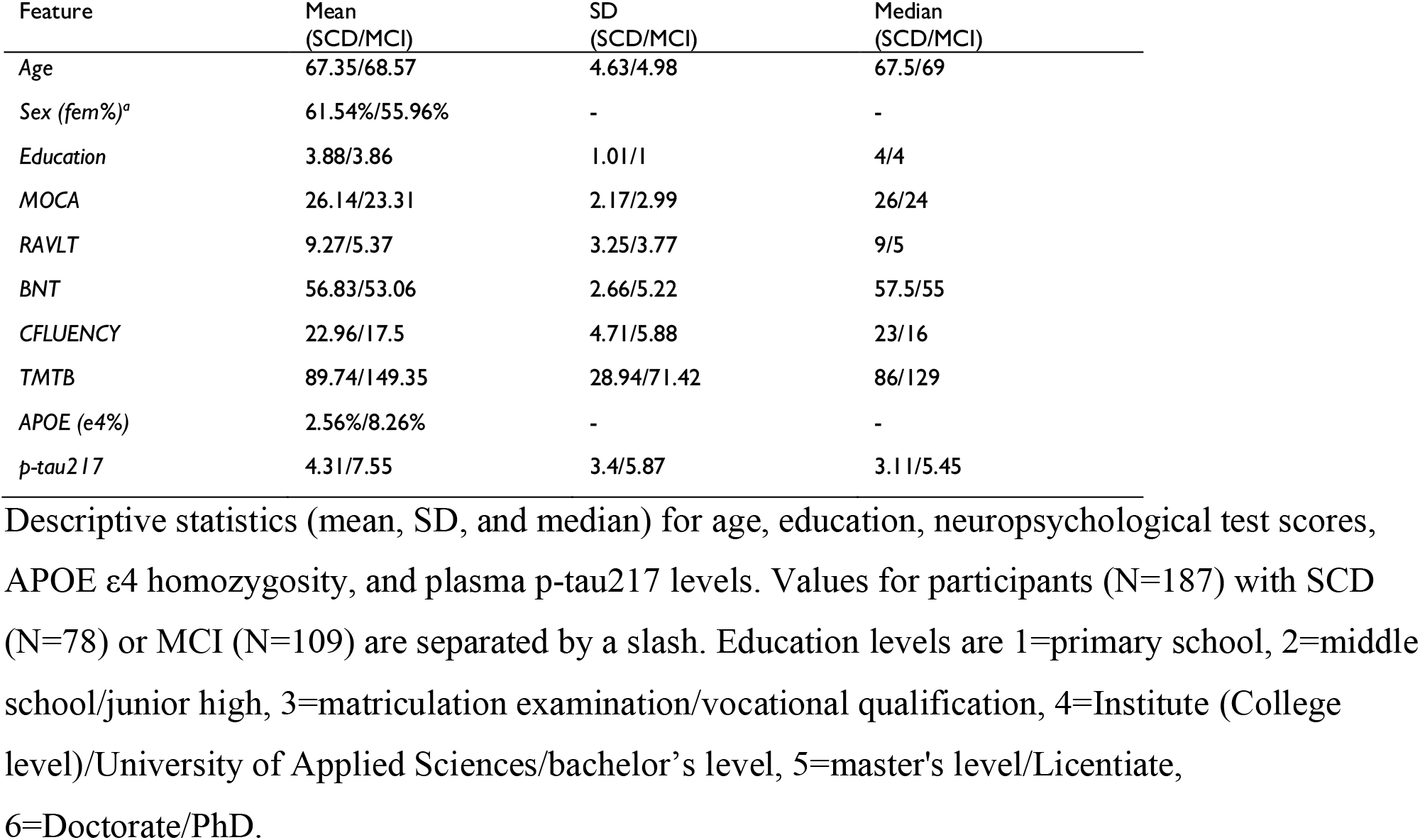
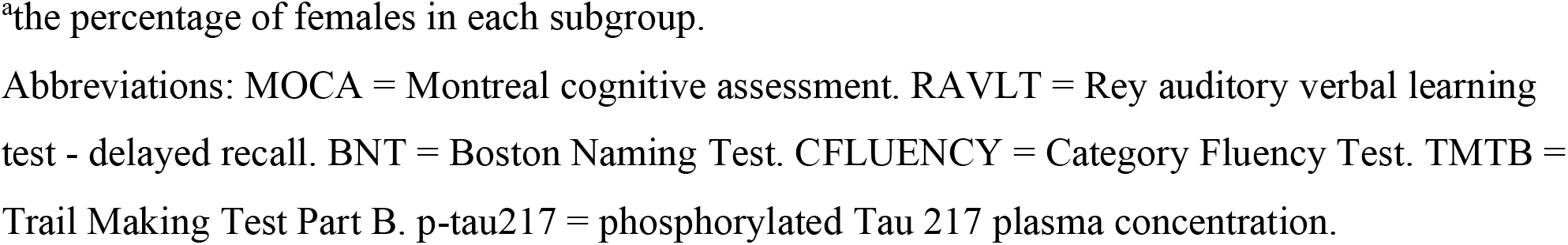
Descriptive statistics.

| Feature | Mean<br>(SCD/MCI) | SD<br>(SCD/MCI) | Median<br>(SCD/MCI) |
| --- | --- | --- | --- |
| Age | 67.35/68.57 | 4.63/4.98 | 67.5/69 |
| Sex (fem%)<sup>a</sup> | 61.54%/55.96% | - | - |
| Education | 3.88/3.86 | 1.01/1 | 4/4 |
| MOCA | 26.14/23.31 | 2.17/2.99 | 26/24 |
| RAVLT | 9.27/5.37 | 3.25/3.77 | 9/5 |
| BNT | 56.83/53.06 | 2.66/5.22 | 57.5/55 |
| CFLUENCY | 22.96/17.5 | 4.71/5.88 | 23/16 |
| TMTB | 89.74/149.35 | 28.94/71.42 | 86/129 |
| APOE (ε4%) | 2.56%/8.26% | - | - |
| p-tau217 | 4.31/7.55 | 3.4/5.87 | 3.11/5.45 |

Descriptive statistics (mean, SD, and median) for age, education, neuropsychological test scores, APOE ε4 homozygosity, and plasma p-tau217 levels. Values for participants (N=187) with SCD (N=78) or MCI (N=109) are separated by a slash. Education levels are 1=primary school, 2=middle school/junior high, 3=matriculation examination/vocational qualification, 4=Institute (College level)/University of Applied Sciences/bachelor's level, 5=master's level/Licentiate, 6=Doctorate/PhD.
<sup>a</sup>the percentage of females in each subgroup.
Abbreviations: MOCA = Montreal cognitive assessment. RAVLT = Rey auditory verbal learning test - delayed recall. BNT = Boston Naming Test. CFLUENCY = Category Fluency Test. TMTB = Trail Making Test Part B. p-tau217 = phosphorylated Tau 217 plasma concentration.

The SCD subgroup comprised individuals who reported memory or cognitive concerns but showed no objective impairment on neuropsychological testing, while the MCI subgroup comprised individuals with objective impairment with activities of daily living largely preserved. Objective cognitive impairment was evaluated using the Consortium to establish a registry for Alzheimer’s disease (CERAD;^37^) battery’s subtasks (covering memory, language, and visuoconstructive domains) and Trail Making Test (TMT) parts A and B (attention/executive domain). Objective decline was defined by at least one cognitive score of ≤1.5 SD below normative values.^3,38,39^ None of the participants met criteria for clinical dementia; global Clinical Dementia Rating (CDR) scores were 0–0.5. Participants with conditions likely to confound cognitive performance, such as significant cerebrovascular disease, heavy alcohol use, or severe psychiatric illness, were excluded from the study.

All participants provided written informed consent to participate in the study. The study received an ethics statement from the Regional Committee of Medical Research Ethics of the Helsinki and Uusimaa Hospital District (HUS), and it was conducted in accordance with The Declaration of Helsinki for experiments involving humans.

### Data acquisition

Blood samples for DNA isolation and plasma protein assessment were collected from all study participants and stored in the Helsinki Biobank (Helsinki Biobank, Helsinki, Finland) for further analysis. Plasma p-tau217 concentrations were measured using the S-PLEX® Human Tau (pT217) Kit (Meso Scale Diagnostics, Rockville, MD, USA) according to the manufacturer’s instructions. EDTA plasma samples were analyzed without dilution using the MSD S-PLEX electrochemiluminescence platform. Briefly, samples and calibrators were added to plates coated with a biotinylated anti-p-tau217 capture antibody, followed by incubation with a TURBO-BOOST® detection antibody, enhancement reagents, and TURBO-TAG® detection solution. Electrochemiluminescence signals were measured using an MSD QuickPlex instrument. Quantification was performed using a seven-point calibration curve generated from recombinant phosphorylated Tau (p-tau217) standards and fitted using a four-parameter logistic (4-PL) regression model with 1/Y² weighting. The assay has a reported lower limit of detection (LLOD) of 880 fg/mL and a lower limit of quantification (LLOQ) of 5,900 fg/mL.

EEG measurements were performed at the BioMag laboratory (HUS Diagnostic Center) using 126 cephalic electrodes prewired in an elastic cap (ANT neuro WaveguardTM), in addition to two auxiliary electrodes for electro-oculogram (EOG) and electrocardiogram (ECG). The ground electrode was placed on the left mastoid. The electrode positions followed the 10-5 system, with the CPz electrode serving as the reference during recordings. Signals were amplified using the eego™ mylab EE-228 amplifier system and digitized for storage with eego™ software, both supplied by ANT Neuro/eemagine Medical Imaging Solutions GmbH (Berlin, Germany). The sampling rate during the recordings was 2000 Hz, with an anti-aliasing filter applied at a cutoff frequency of 520 Hz (Haraldsen et al., 2024).

All participants underwent resting-state EEG measurements (wakeful rest) acquired in four alternating 5-min runs of eyes-open (EO) and eyes-closed (EC) conditions at the initial time point (*n* = 187), and again eight months later (*n* = 173). For the present analyses, only EC data were used.

In addition, the participants were evaluated neuropsychologically in two separate sessions (on average 14,0±13,3 SD days apart): first with the Montreal Cognitive Assessment (MOCA) and CERAD battery for characterization of global cognitive performance and MCI status and, second, with a broader set of neuropsychological tests (see Haraldsen et al.^40^ for the full neuropsychological protocol) from which the most pivotal, domain-selective variables were derived for the present analysis. The chosen neuropsychological scores included the delayed (20 min) word list recall of the Rey Auditory Verbal Learning Test – delayed recall (RAVLT) for the memory domain; the part B of the Trail Making Test (TMTB) for the attention/executive domain; and the Boston Naming Test (BNT) and verbal category fluency (CFLUENCY) for the language domain. This core set of test scores has been shown to reliably capture key cognitive deficits ^6,41-43^ and to align with empirically derived neuropsychological subtypes in MCI and AD.^44,45^

### Data preprocessing

#### EEG data preprocessing

MNE Python (version 1.7) was used for most EEG data preprocessing.^46^ Raw EEG data were first converted to a standardized brain imaging data structure (BIDS) format.^47^ Power-line noise at 50 Hz was removed using the Zapline plus algorithm in EEGLAB.^48^ Bad channels were identified using the pyprep software for EEG preprocessing and manual inspection.^49^ Ocular, cardiac and prominent muscle artifacts were removed using independent component analysis (ICA) using the FastICA algorithm as implemented in MNE Python; the results were manually confirmed. Remaining muscle artifacts were manually annotated, and the contaminated data segments were discarded.

Power spectral density (PSD) was estimated over the 5+5 min EC time segments for each EEG channel from 1 to 90 Hz using Welch’s method with an 8192-point Fast Fourier Transform, 0% overlap, and Hamming windowing. The PSDs were converted to a decibel scale (dB) to emphasize minor amplitude differences in the data. Frequency bins affected by the 50 Hz power-line interference were discarded. The FOOOF algorithm (version 1.1.1) was used to separate the spectra into aperiodic and periodic components.^50^ The power spectra were parameterized over the frequency range from 2 to 90 Hz, with a maximum of 12 peaks and a minimum peak height of 0.1 to reduce the detection of spurious spectral peaks.

For the periodic components, average absolute power and mean peak frequencies were calculated for five regions of interest (ROIs: occipital, fronto-central, occipital-extended, fronto-central-left, fronto-central-right; see **Fig. 1**) at two frequency bands (7–13 Hz [alpha] and 13–30 Hz [beta]), selected to capture the main spatial distribution of most prominent oscillatory activity. As the occipital peak frequencies have been shown to decrease in later stages of dementia, a broad alpha band range was used.^51^ For the aperiodic component, we extracted the 1/f slope and offset for each ROI. In total, we derived 30 sensor-level EEG features: 20 periodic (5 ROIs × 2 frequency bands × 2 features [absolute power and peak frequency]) and 10 aperiodic (5 ROIs x 2 features [1/f slope and offset]).

**Figure 1.**
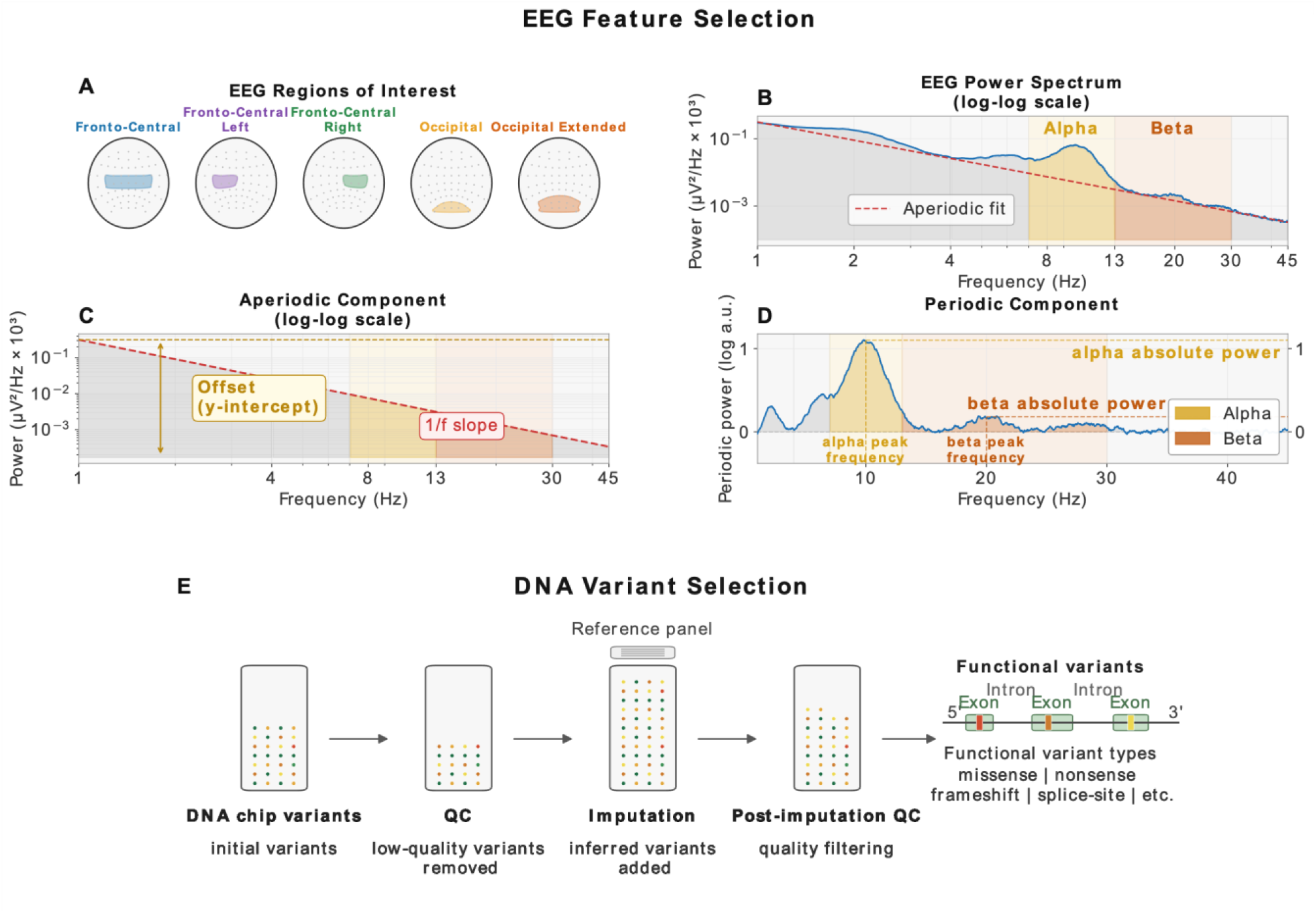
EEG-feature and DNA-variant selection. **(A)** Features were averaged over targeted regions of interest (ROI). **(B)** EEG power spectral density (PSD) was estimated using Welch’s method with the Fast Fourier Transform, and aperiodic and periodic components were extracted using the FOOOF algorithm. EEG features consist of **(C)** two aperiodic features (1/f offset and slope) and **(D)** average spectral power and peak frequency at 7-13 Hz and 13-30 Hz frequency bands. **(E)** In DNA variant selection, raw DNA chip variants were preprocessed through quality control (QC), imputation, post-imputation QC and then filtered by selecting only functional variants.

#### Genotype data preprocessing

Genomic DNA was isolated from the blood samples of study participants with consent for Helsinki Biobank (*n*=187) using the QIAsymphony DSP DNA Midi kit (QIAGE, Hilden, Germany, catalog number: 937255) with a QIAsymphony platform according to the manufacturer’s instructions. DNA quantity was measured using Qubit or Nanodrop (Thermo Fisher). Genotyping was performed according to the manufacturer’s instructions (Illumina Inc., San Diego, CA, USA). The DNA array was Illumina’s InfiniumTM Global Screening Array MD (GSAMD-24v3), with 730059 SNPs, or with 730059 SNPs + 30000 custom SNPs targeting Finnish heritage.

All genotypes were called using GenomeStudio v. 2.0.4 software, and SNPs were manually checked if they failed quality control criteria due to low call rates, bad cluster separation (<0.3), low signal intensity, quality scores, and heterozygote excess.

Standard quality control was performed using PLINK v.1.9 or v2.0.^52^ Sex inconsistencies and non-random missingness in variants by haplotype were checked, requiring minimum variant call rate >95%, minimum sample call rate >90%, Hardy-Weinberg equilibrium P-values > 0.00001, and a minor allele frequency of > 0.1%. Ambiguous strand variants (A/T and G/C) were removed. Relatedness to the 3rd generation (Identity by Descent, IBD > 0.185) was tested, and from the detected relative pairs, only one sample was randomly selected for subsequent analysis. Heterozygous outliers deviating more than 3 SD from the mean observed heterozygosity rate were removed. European ancestry was confirmed with principal component or multidimensional scaling analyses. In total, 693,317 variants and 184 individuals passed the quality control before imputation.

Pre-phasing and imputation to a standard reference panel from the 1000 Genomes Project (October 2020 release, aligned to the GRCh38 reference genome), were performed using the Eagle v.2.4.1 and Beagle v.5.4 programs.^53,54^ Imputed variants were filtered using a dosage R-squared (DR2) value > 0.5. In total, 184 individuals (59 % females) and 14,716,210 variants passed quality control.

Gene regions (in total 17,735) were identified using ANNOVAR software (version 2017July17).^55^ For each gene, only functional variants (nonsynonymous, synonymous, frameshift deletion, frameshift insertion, non-frameshift deletion, non-frameshift insertion, splicing, stop gain, and stop loss) were selected for the analysis, and all variants with zero variance were removed. The statistical analysis was performed using 16,935 genes and 90,607 functional variants.

#### Data analysis

Of the 187 participants at the baseline, 173 completed the 8-month follow-up EEG measurement. After preprocessing, EEG data of 170 participants qualified for merging with covariate and genotype data, leaving in total the data of 169 participants for BRRR analysis.

Gene-level associations in relation to the periodic and aperiodic EEG features (see above) were analyzed using resting-state EC (5+5 min) data from two measurement time points (initial time point and a repeated measurement 8 months later). The covariates were age, sex, 20 principal components of the imputed genotype, information of the measurement session (two runs in two visits), and functional variants of each gene. After EEG and genotype data preprocessing and merging, data from 169 individuals were included in the statistical analyses. Bayesian reduced rank regression (BRRR) was first applied for dimensionality reduction and regression, yielding ten and five latent EEG features for periodic and aperiodic features, respectively.^56^ Subsequently, using unsupervised k-means clustering (R 4.5.1; stats 4.5.1), literature review of neurobiological relevance, and comparison against known AD genes were performed for the candidate gene sets that passed specific significance level checks. A schematic overview of the analysis is presented in **Fig. 2**.

**Figure 2.**
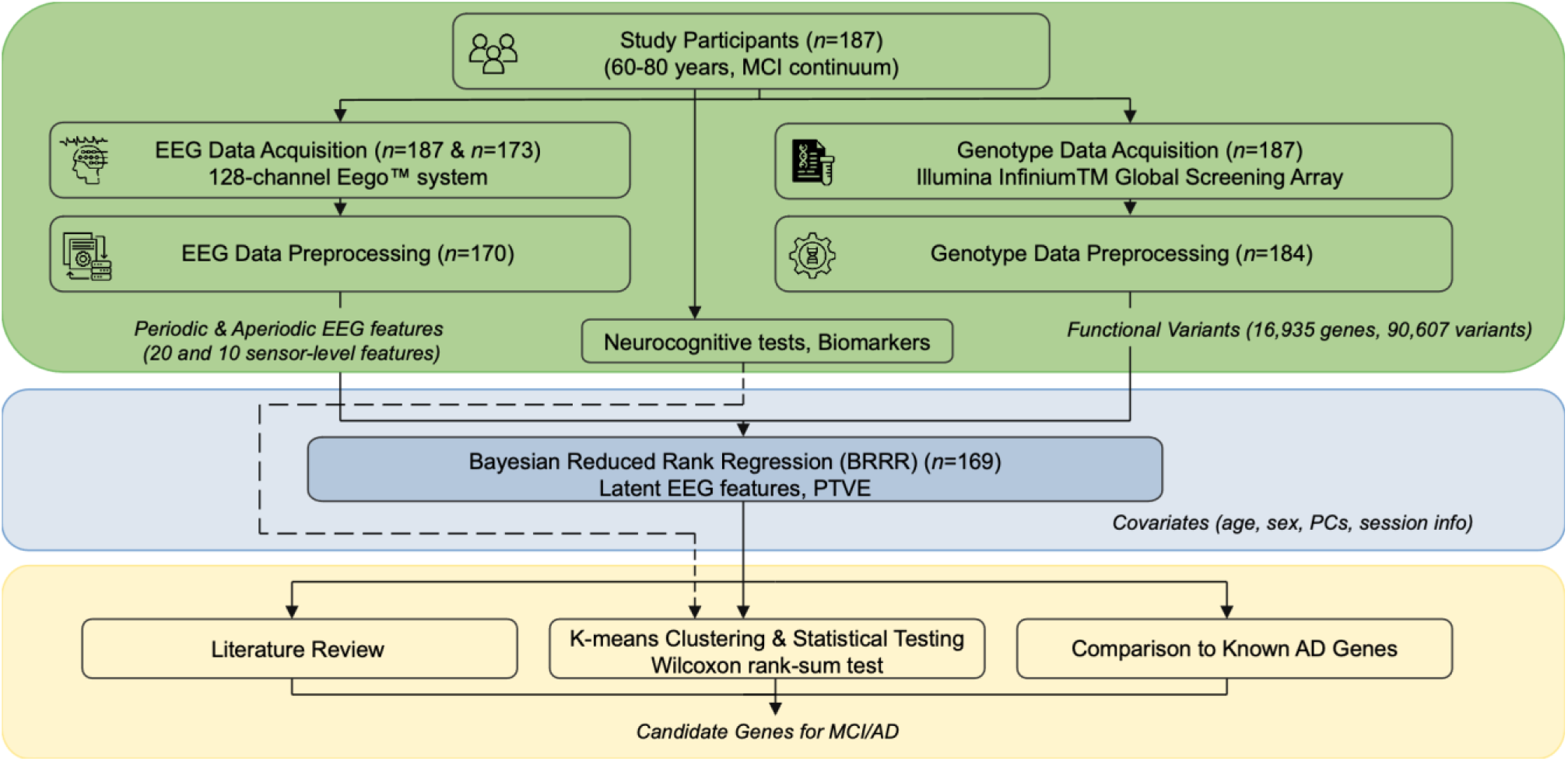
Schematic overview of the analysis integrating EEG and genomic data to identify candidate genes associated with MCI. **Data Acquisition and Preprocessing (green)**: Resting-state EEG recordings (*n* = 187 at an initial time point and *n*=173 at 8 months) were processed to extract periodic and aperiodic power spectral density (PSD) features. Concurrently, genotype data (*n* = 187, *n* = 184 after quality control) underwent imputation to the 1000 Genomes reference panel, and functional annotation. **Statistical Modeling (blue)**: Associations between functional gene variants and EEG phenotypes (*n*=169) were evaluated using Bayesian Reduced Rank Regression (BRRR).

The model included age, sex, session info and genetic principal components as covariates. **Post-hoc Analysis (yellow)**: Identified genes were characterized through literature review of neurobiological relevance, by comparison with known AD risk loci, and unsupervised K-means clustering of participants to assess differences in clinical biomarkers. Abbreviations: ICA, Independent Component Analysis; PTVE, Proportion of Total Variance Explained.

##### Bayesian reduced rank regression analysis

Bayesian reduced rank regression (BRRR) was used to reduce the dimensions of the dependent variables and to identify genes associated with these latent EEG features.^56^ After excluding variants with zero variance from the data, 16,935 gene regions remained. The BRRR model is defined as

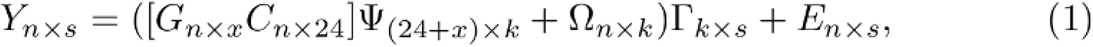

where **Y**_n*s_ contains *n* samples (here 169 participants) and *s* EEG-features (here 20 for periodic data and 10 for aperiodic data). **G**_n*x_ contains the x functional variants of the gene in question. **C**_n*24_ contains covariates, including session information (two runs in two visits), age, sex, and the first 20 principal components of imputed genotype data. **Ψ_(24+x)*k_** is a low-dimensional regression coefficient matrix, where **k** is the rank of the model and defines the number of latent factors determined. **Ω**_n*k_ contains unknown factors representing noise in the latent space. **Γ**_k*s_ is a projection of the latent space to the EEG feature space. The product of **ΨΓ** is a standard regression coefficient matrix **β** with a rank of **k**. **Ε**_n*s_ describes residual noise in the observation space.

The explanatory power of each gene in the EEG feature space was measured as the model’s proportion of the total variance explained (PTVE) in **Y**. PTVE is, defined as

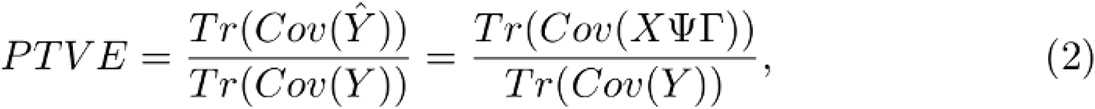

where **Tr** is the trace operation, **X =** [**G C]** and **Cov** refers to the covariance matrix. The relative PTVE was calculated by subtracting the PTVE of the full model from that of the null model containing covariates without gene variants.

EEG features were min–max scaled to the range [0,1], and gene data and other covariates were z-score scaled to have a zero mean and unit variance. The latent noise variance prior was empirically set to 0.01, and the model rank **k** to 10 for periodic data and 5 for aperiodic data. Shrinkage parameters a1 and a2 were set to 2, and the local gamma shrinkage parameter was set to 1. The BRRR model was initialized using the first **k** principal components of the **Y** matrix, and the parameters were trained with 2000 iterations for periodic data and 1500 iterations for aperiodic data, using the first 50% of the samples as the burn-in period. BRRR model was trained for each gene separately.

Convergence and stability checks were performed using Gelman–Rubin statistic (R-hat), with values summarized across all genes; the effective sample size (ESS) was estimated for both periodic and aperiodic data sets. R-hat was an average of 1,003±0.008 and 1,004±0.01, respectively, indicating no obvious bias between chains. The average bulk ESS was 197±34/151±33, while the corresponding ESS for the tail was 174±33/132±28. These values suggest sufficient mixing of the chains and effective sampling across posterior distributions, indicating stable performance in line with earlier recommendations.^56^

To estimate the significance of the results, a two-stage label permutation was performed by randomly shuffling the participant labels in the EEG feature data for a random baseline. First, 500 label-permuted runs were performed for each of the 16,935 genes. P values were calculated, and the most significant genes were subjected to additional permutations for more precise estimates.

Accordingly, 1802 periodic and 1507 aperiodic genes (both P < 0.05) underwent a second round of label permutation with 800 additional runs. The genomic inflation factors (λ) were 0.997 for periodic data and 0.237 for aperiodic data. QQ plots are shown in **Supplementary Figs S1** and **S2.**

#### Genotype clustering and phenotypic profiling

To examine whether candidate genes associated with periodic and aperiodic EEG features also relate to cognitive or biological characteristics of the subjects, we performed unsupervised k-means clustering on the periodic and aperiodic candidate gene sets. Clusters with fewer than three samples were removed. The Wilcoxon rank-sum test was used to assess the differences between the clusters. The Benjamini–Hochberg procedure was used to adjust for multiple comparisons. ANOVA was used to statistically identify and sort the top 50 variants that were most important for creating these clusters. Based on this sorted variant list, the corresponding top five genes most important for cluster formation were selected, and the clustering and statistical testing of features were rerun.

### Literature review

A literature review (with “neurobiology”, “brain” and “neurodegeneration” as search words) was conducted on the periodic and aperiodic candidate gene sets with *P* < 0.00077 to indicate that the identified genes have neurobiological functions. In total, 32 periodic and 20 aperiodic candidate genes were identified.

## Results

### EEG-associated genes include AD-associated loci and genes linked to neuronal function

Several promising candidate genes were identified with BRRR analyses at significance thresholds (uncorrected, *P* < 0.00077), but these did not survive correction for multiple testing, likely reflecting limited statistical power given the current sample size (*n*=169). A full gene lists, gene function literature review with references and variant annotations are provided in **Supplementary Tables S1–S3 and Supplementary Text.**

In total 145 genes associated with periodic EEG features and 39 with aperiodic features (*P* < 0.005). Two candidate genes, RASGEF1C and TREML2, fall within loci previously associated with Alzheimer’s disease (AD).^22^ The previously reported variants at these loci are not in linkage disequilibrium with the functional variants analyzed here, indicating they likely do not tag the same causal signal. One candidate gene, C5AR2, is in a locus associated with AD in the Finnish genome and health database FinnGen (phenotype “AD WIDE” in data release 12; P < 1e-8; ∼200 genes;^57^).

Out of 32 top candidate genes associated with periodic EEG features (*P* < 0.00077), 9 (28.1%) were implicated in synaptic transmission and neuronal excitability, and 12 (37.5%) in neurodevelopmental processes (neuronal differentiation and structure). Among 20 top candidate genes associated with aperiodic EEG features (*P* < 0.00077), the corresponding counts were 5 (25%) and 7 (35%). Representative genes include FRRS1L, SPARCL1, EGR1, JPH3, IRAG2, EMC7, MEG9, KIAA0513, and ADAR.

### Comparison of periodic and aperiodic EEG candidate genes reveals distinct biological profiles

Comparison of the top genes associated with periodic and aperiodic EEG features revealed distinct functional enrichment profiles. Periodic candidate genes were more frequently annotated to intracellular neuronal maintenance than aperiodic ones (34.4% vs. 15%). Representative periodic candidate genes included SCP2, JPH3, LMX1B, USP1, ZYG11A, and GCC2.

In contrast, aperiodic candidate genes showed greater involvement in neuroinflammation compared to the periodic ones (30% vs. 18.8%). Representative aperiodic candidate genes included C5AR2, EMB, NCF1, PHLDA1, SCRG1, and TREML2. Functional category comparisons between genes that were associated with periodic vs. aperiodic EEG features are provided in **Supplementary Table S4 and Supplementary Text**.

### Variant clustering reveals MCI subgroups differing in p-tau217 and delayed recall

We performed unsupervised clustering of EEG-associated candidate genes (periodic *P* < 0.005, *n* = 145; aperiodic *P* < 0.005, *n* = 39), yielding two clusters. We visualized the clusters with selected cognitive and biomarker features and tested for between-cluster differences. For the set of genes associated with periodic features, the participant clusters differed statistically significantly for their plasma p-tau217 and delayed recall (RAVLT) scores (Benjamini–Hochberg *P* = 0.02; **Fig. 3, Table 1, Supplementary Table S5-S7**). The top five genes most important for cluster formation (ACAN, INPP5B, CAMKK2, CABIN1, SPATA13; 80 functional variants) reproduced this phenotypic pattern but did not reach significance for p-tau217 or delayed recall after multiple testing (**Fig. 3**). For the set of genes associated with aperiodic features, the results were non-significant (**Supplementary Table S5**).

**Figure 3.**
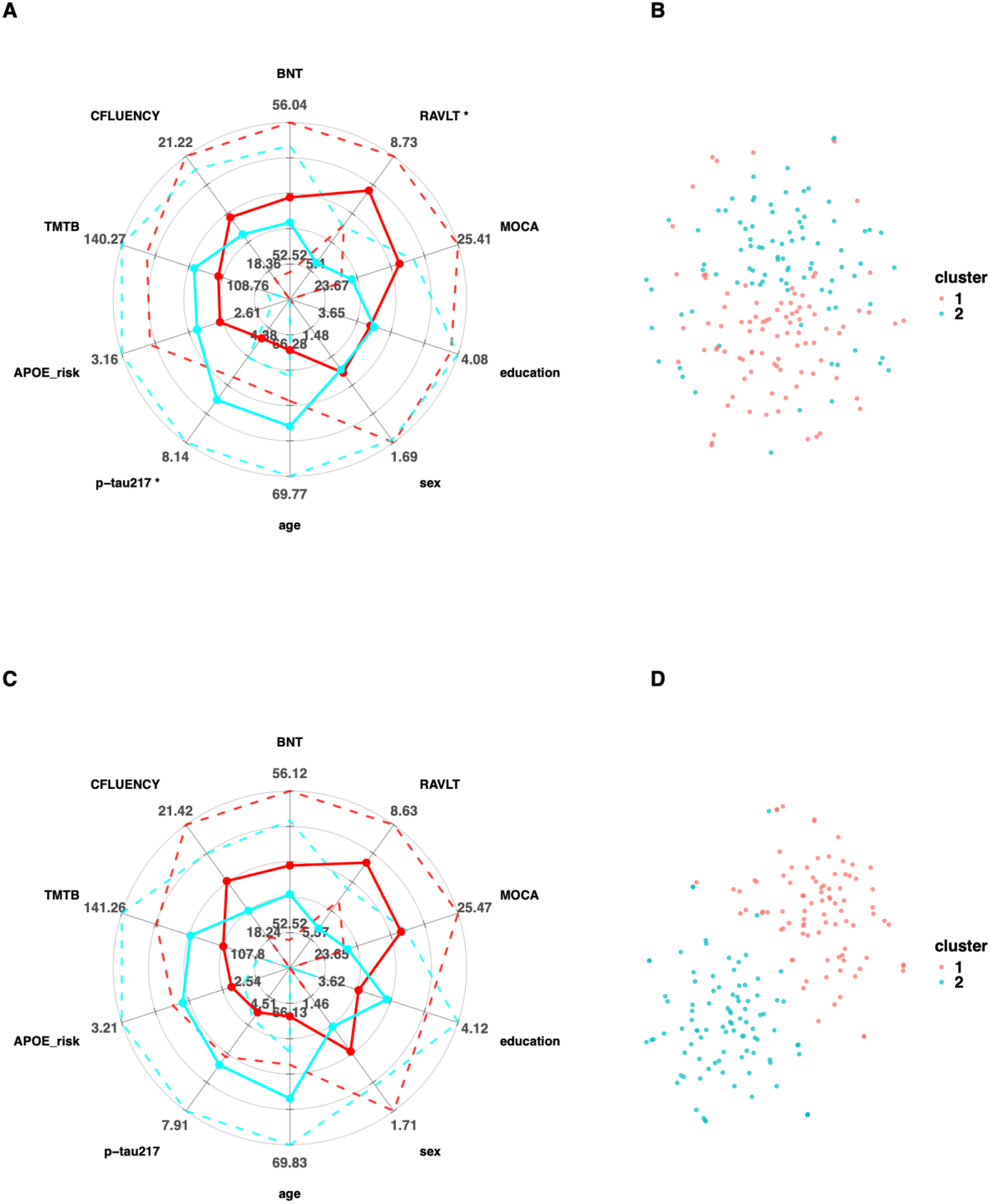
Phenotypic profiles of genetically distinct subject clusters. (**A)** Radar chart illustrating the multimodal characterization of the subject clusters (*k* = 2) identified via K-means clustering of genetic variants in selected genes associated with periodic EEG features (*P* < 0.005, *n* = 145). Each radial axis represents a demographic, clinical, or biomarker feature, scaled from the minimum (center) to the maximum (outer edge) value observed in the cohort. Profiles show the mean value for each cluster (solid red and blue lines). Dashed lines indicate 95% confidence intervals.

Phenotypic feature variation reveals distinct clinical signatures associated with the genetic subgroups; notably, statistical testing identified significant differences between clusters in plasma p-tau217 levels and RAVLT scores (*P* = 0.02, Benjamini-Hochberg corrected). (**B)** Two-dimensional visualization (t-SNE) of the K-means clusters based on the genetic variants in selected periodic genes (*P* < 0.005, *n* = 145). (**C)** Radar chart displaying the phenotypic characteristics of clusters re-identified using five genes with the most discriminative power. Features were not statistically significant between clusters. (**D)** Two-dimensional visualization (t-SNE) of the K-means clusters based on the five top discriminative genes. Abbreviations: APOE_risk = estimated risk of developing dementia based on APOE allele (range 1-5; 5=highest risk). MOCA = Montreal cognitive assessment (higher value = better performance). RAVLT = Rey auditory verbal learning test - delayed recall (higher value = better performance). BNT = Boston Naming Test. CFLUENCY = Category Fluency Test. TMTB = Trail Making Test Part B. p-tau217 = phosphorylated Tau 217 plasma concentration. Education: higher value = higher level of education. SEX: 1 = male, 2 = female. \**P* ≤ 0.05, Benjamini-Hochberg corrected

**Table 2.** Between-cluster comparison of key features after k-means clustering on periodic gene variants.

| Feature | F Statistic | P Value | P Adjusted |
| --- | --- | --- | --- |
| age | 3443 | 0.038 | 0.126 |
| sex | 4205 | 0.952 | 0.952 |
| education | 4156 | 0.932 | 0.952 |
| MOCA | 4558 | 0.297 | 0.717 |
| RAVLT | 5175 | 0.006 | **0.028\*** |
| BNT | 4411.5 | 0.367 | 0.717 |
| CFLUENCY | 4344 | 0.660 | 0.881 |
| TMTB | 3903 | 0.430 | 0.717 |
| APOE_risk | 4063.5 | 0.705 | 0.881 |
| p-tau217 | 3145.5 | 0.004 | **0.028\*** |

P values are based on the Wilcoxon test; Benjamini-Hochberg was used to correct for multiple tests.
Abbreviations: APOE\_risk = risk of developing dementia based on APOE allele (range 1–5).
MOCA = Montreal cognitive assessment. RAVLT = Rey auditory verbal learning test - delayed recall. BNT = Boston Naming Test. CFLUENCY = Category Fluency Test. TMTB = Trail Making Test Part B. p-tau217 = phosphorylated Tau 217 plasma concentration.
\* $P \leq 0.05$

## Discussion

Our study aimed to integrate resting-state EEG with genome-wide functional variation to probe the genetic architecture of EEG-derived endophenotypes along the mild cognitive impairment (MCI) continuum, and to link these intermediate traits to cognition and plasma biomarkers. Using Bayesian reduced rank regression (BRRR), we identified candidate genes associated with periodic and aperiodic EEG features, with convergent evidence for roles in synaptic function, excitability, and neurodevelopment. Unsupervised clustering based on variants associated with the periodic EEG features delineated two subgroups within the MCI continuum that differed in their plasma p-tau217 and delayed verbal recall (RAVLT) scores, suggesting partially dissociable processes of tau-related pathology and network mechanisms supporting episodic memory that need not progress in lockstep.

The genetic landscape uncovered here largely falls outside established Alzheimer’s disease (AD) GWAS signals,^22^ with the exceptions of TREML2 and RASGEF1C loci and C5AR2 residing within a FinnGen AD-associated region. This divergence is expected given our trait-focused approach based on functional variants and latent EEG phenotypes rather than clinical diagnosis. The structural and functional connectome of the brain itself is strongly influenced by genetics: At least 6,443 independent genome-wide loci have been found to associate with MRI derived brain imaging phenotypes that reflect brain structure, function, and connectivity.^58^ The concentration of our candidate genes in synaptic assembly, receptor biogenesis, calcium signaling, and intracellular maintenance supports the biological relevance of these EEG-derived traits as readouts of network integrity and excitability.

The identified loci suggest complementary mechanisms impacting EEG features and AD-relevant biology. RASGEF1C belongs to the Ras guanine nucleotide exchange factor family expressed in the brain (Human Protein Atlas; ^59^) and implicated in synaptic plasticity,^60,61^ plausibly contributing to EEG periodic traits via synaptic signaling mechanisms. TREML2, a myeloid receptor with pro-inflammatory effects^62,63^ carries rs3747742 that has previously been linked to reduced AD risk and white matter hyperintensity burden.^64^ C5AR2 is a complement receptor with neuroprotective properties that attenuates C5AR1-mediated proinflammatory signaling.^65-68^ Together, these observations support contributions from synaptic signaling, microglial/immune function, and complement pathways.

Periodic and aperiodic EEG features mapped onto partially distinct processes. Periodic features (alpha/beta power and peak frequency) yielded candidate genes favoring intracellular neuronal maintenance and signaling (including lipid handling, ER–lysosomal trafficking, and ubiquitin-mediated turnover; e.g., SCP2, JPH3, LMX1B, USP1, GCC2), consistent with the dependence of large-scale alpha/beta oscillations on synaptic excitation–inhibition (E – I) dynamics and neuro-energetic constraints.^69-73^ Instead, biophysical modeling and earlier empirical data have linked the aperiodic (1/f-like) spectral exponent (slope) to circuit-level E–I balance, and human studies have shown robust modulation of the aperiodic slope by arousal and sleep depth,^31,74,75^ whereas the offset primarily reflects broadband power level and tracks population spiking.^76,77^ Neuroimmune signaling can directly influence both processes: microglial and astrocytic cytokines and complement pathways can modulate synaptic strength and pruning, thereby shifting E–I balance, and inflammatory cytokines may alter arousal/sleep regulation.^78-81^ In this context, our candidate genes associated with aperiodic EEG features, enriched for neuroinflammatory pathways (e.g., TREML2, C5AR2, NCF1, PHLDA1, EMB, SCRG1), provide a plausible link between immune tone and EEG dynamics.

Two key indices of AD-type neurodegeneration, p-tau217 and delayed recall (RAVLT), differed significantly across the periodic gene-based clusters. Although both measures are closely linked to AD pathology, they likely reflect partly overlapping biological processes operating at different levels of the disease cascade. Plasma p-tau217 reflects a defect in tau phosphorylation system and postsynaptic pathology related to amyloid-β burden^82^, whereas delayed recall relies on hippocampal–medial temporal network function modulated by synaptic plasticity, inhibitory tone, and extracellular matrix scaffolding. EEG preferentially tracks neocortical dynamics, whereas tau accumulation and episodic memory depend critically on medial temporal circuitry. Nevertheless, hippocampal-cortical interactions contribute substantially to large-scale cortical oscillations, linking scalp-recorded activity to episodic memory networks. The medial temporal lobe (MTL)-related memory network extends beyond the medial temporal structures and involves distributed cortical regions, particularly the precuneus and anterior cingulate cortex, which constitute the key nodes of the default mode network (DMN). As alpha- and beta-band oscillations are prominent signatures of large-scale DMN dynamics, these rhythms may partly reflect episodic memory functions supported by coordinated MTL-cortical interactions. In this framework, the EEG measures examined here can be viewed as downstream manifestations of activity within episodic memory networks, providing a plausible biological basis for the observed associations between periodic EEG-associated genetic variation and delayed recall performance. Nevertheless, synaptic loss has been documented in the neocortex of individuals with MCI^83^ and decreasing functional connectivity between cortex and medial temporal circuits may reflect preclinical Alzheimer’s disease.^84^

The five most discriminant genes—ACAN (perineuronal nets), INPP5B (phosphoinositide signaling), CAMKK2 (Ca2+-dependent signaling, tau phosphorylation), CABIN1 (calcineurin pathway), and SPATA13 (spine/axon development)—map onto these network-supporting mechanisms^85-95^ that could sustain memory performance despite tau burden, or conversely, confer vulnerability to memory decline without yet marked changes in p-tau217. Notably, MAPT responsible for encoding the tau protein did not appear among the top periodic genes, and among tau-related kinases only CAMKK2 featured prominently,^89^ further supporting the view that network-level support can vary relatively independently of canonical tau loci.

Phenotypically, the cluster separation was specific to p-tau217 and RAVLT, not to MoCA, TMT-B, CFLUENCY, or BNT, suggesting selectivity related to episodic memory domain. Among cognitive measures, RAVLT is one of the most effective, early indices in identifying MCI and AD^42,96,97^ and, unlike executive measures like TMT-B,^43^ it specifically captures the amnestic profile characteristic of typical AD.^4,6,98^ P-tau217 and RAVLT are correlated overall,^99^ and both have been linked to core AD pathology, including medial temporal atrophy,^100-102^ altered amyloid β/tau levels in the cerebrospinal fluid,^103-106^ and positive amyloid β/tau positron emission tomography (PET) findings in the brain.^102,107-109^ Moreover, both predict conversion from MCI to dementia with relatively high discriminative accuracies (AUC ≈ ∼0.75–0.9; ^6,110^). Despite these associations, longitudinal studies suggest that plasma p-tau217/181 and cognitive measures, including RAVLT, may follow partially decoupled and nonlinear trajectories, with biomarker changes preceding and only inconsistently tracking concurrent cognitive decline.^111-113^ The present cross-sectional study further highlights dissociation of these different classes of measures at the primary level of neurophysiology and underlying genes. Notably, EEG/MEG spectral and network features have also been associated with neural reserve—e.g., preserved alpha synchrony, efficient large-scale connectivity, and favorable E–I balance^50,114,115^—which can support memory performance despite underlying genetic and other biological risks, offering a plausible substrate for the observed divergence.

Limitations in this study should be acknowledged. First, none of the identified genetic associations survived multiple test corrections, and therefore the reported candidate genes should be regarded as hypothesis-generating. Although the study leveraged multimodal data including longitudinal EEG recordings, cognitive assessments, plasma biomarkers, and genome-wide genotype information, the sample size remained modest for a genetic association study with likely limited statistical power. Second, our approach focused on functional variants aggregated at the gene level and therefore did not capture the full spectrum of genetic influences on EEG phenotypes. Third, the clustering analyses were exploratory and performed within the same dataset used for gene discovery, warranting cautious interpretation until independent replication is available. Finally, the analyses were based on sensor-level EEG measures from a relatively homogeneous Finnish cohort spanning the MCI continuum, and the generalizability of the findings to other populations and to source-resolved electrophysiological measures remains to be established. Despite these limitations, the convergence of multiple lines of evidence—including overlap with AD-associated loci, biologically plausible functional annotations, and associations with p-tau217 and delayed verbal recall— suggests that the identified candidate genes capture biologically meaningful variation relevant to cognitive decline and AD-related pathology.

In conclusion, genetically anchored EEG signatures delineate complementary biological dimensions of MCI continuum—tau-related biochemical pathology and network resilience supporting episodic memory—that are only partially coupled. Replication studies, longer follow-up, studies linking aperiodic EEG-associated genes to cognitive biomarkers, and source-resolved EEG/MEG analyses will be essential to validate these results and to test whether they improve risk stratification and guide mechanism-informed interventions in MCI at risk of AD.

## Data availability

According to restrictions imposed by our research ethics committee, and following the current Finnish regulations, we are not allowed to share raw brain imaging data openly. However, the individual models derived from the data can be shared, and code used for the machine learning analysis is available at BioMag Laboratory GitLab repository (https://version.aalto.fi/gitlab/biomag-pipelines/brrr_mci_genes.git).

