## Supplementary material for "Genetic associations with EEG signatures in mild cognitive impairment: insights into Alzheimer’s disease pathophysiology"

#### Table of Contents

SUPPLEMENTARY MATERIAL 1
- Supplementary Figure S1 QQ-plot of relative PTVE values with periodic data. 2
- Supplementary Figure S2 QQ-plot of relative PTVE values with aperiodic data. 3
- Supplementary Table S1 Literature search for top periodic candidate gene set (p < 0.00077, n=32) 4
  - References to literature search for top periodic candidate gene set (p < 0.00077, n=32) 5
- Supplementary Table S2 Literature search for top aperiodic candidate gene set (p < 0.00077, n=20) 10
  - References for literature search of the top aperiodic candidate gene set (p < 0.00077, n=20) 11
- Supplementary Table S3 Descriptive statistics of AD associated genes and variants 14
- Supplementary Table S4 Top (P < 0.00077) Periodic And Aperiodic Gene Characterization 15
- Detailed Gene Classification 15
  - 1. Synaptic Transmission & Excitability 15
  - 2. Neuronal Differentiation & Structure 16
  - 3. Metabolism, ER Stress & Homeostasis 16
  - 4. Neuroinflammation & Immune Regulation 16
  - 5. Gene Regulation & Epigenetics 16
  - 6. Vascular / Other 17
- Supplementary Table S5 Between-cluster comparison of key features after k-means clustering on aperiodic gene variants. 17
- Supplementary Table S6 Top periodic candidate gene set (p < 0.0046, n=145) 18
- Supplementary Table S7 Top aperiodic candidate gene set (p < 0.0046, n=39) 19
- Literature review of the candidate genes 20
- Literature review references 22

**Supplementary Figure S1 QQ-plot of relative PTVE values with periodic data.**

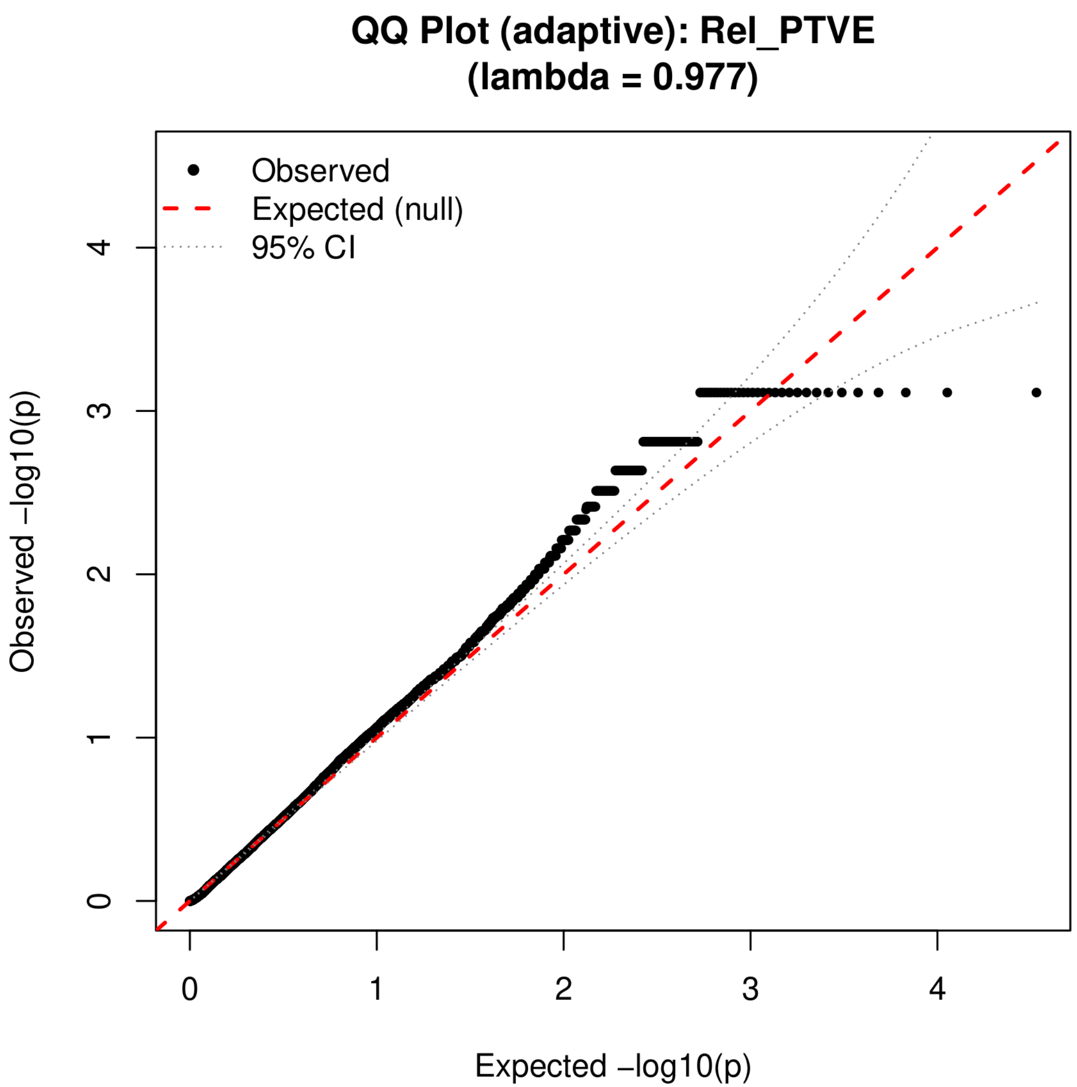

**Supplementary Figure S2 QQ-plot of relative PTVE values with aperiodic data.**

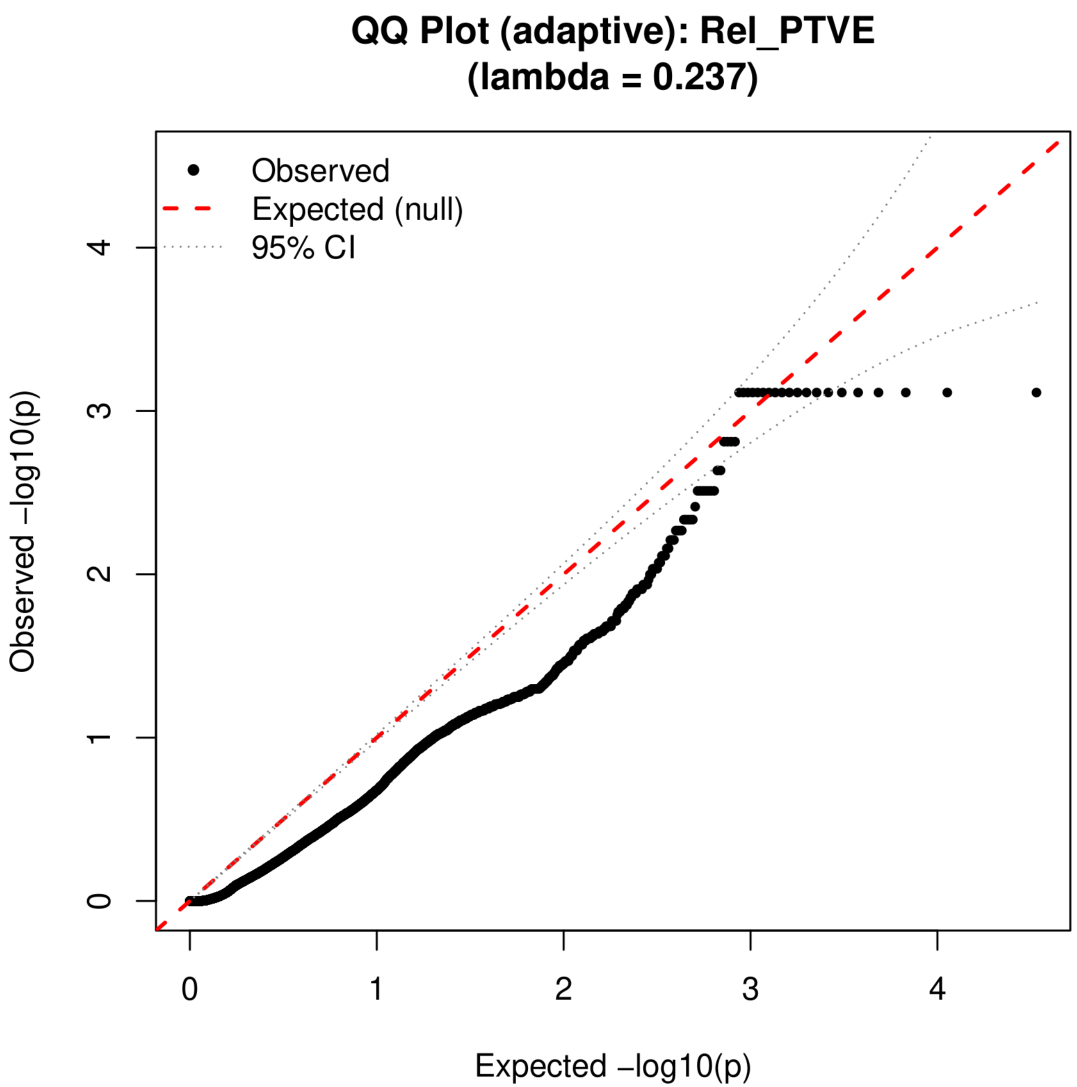

#### Supplementary Table S1 Literature search for top periodic candidate gene set (p < 0.00077, n=32)

| Gene | Association | Citation |
|---|---|---|
| SPARCL1 | Astrocyte-secreted Hevin promotes formation of structurally mature excitatory synapses; biomarker for AD | Kucukdereli et al 2011; Kärkkäinen et al 2025 |
| FRRS1L | FRRS1L is required for AMPA receptor complex assembly – receptor in glutamatergic neurotransmission; loss causes epileptic-dyskinetic encephalopathy; candidate gene in autism | Stewart et al 2019; Madeo et al 2016; Lee et al 2025 |
| KMT2C | H3K4 methyltransferase, involved in neurodevelopmental diseases via epigenetic process. Implicated function in learning and memory | Koemans et al 2017 |
| LMX1B | required for the normal execution of the autophagic-lysosomal pathway and for maintaining the integrity of dopaminergic nerve terminals; essential for specification and survival of central serotonergic neurons; essential for survival of dopaminergic neurons. | Ding et al 2003; Laguna et al 2015 |
| EGR1 | EGR1/Zif268 required for late-phase LTP and consolidation of long-term memory; regulates the expression of acetylcholinesterase; implicated a role in AD | Jones et al 2001; Hu et al 2019 |
| ADAR | involvement in RNA editing (catalyzes adenosine to inosine), neuronal homeostasis, synaptic plasticity, and innate immune regulation. Implicated in neurodegeneration. | Cheng et al 2025 |
| TREML2 | modulates inflammation by regulating microglial polarization and NLRP3 inflammasome activation; Protective TREML2 missense variant associates with reduced Alzheimer's disease risk; associates with whitematter hyperintensity volume | Wang et al 2023; Benitez et al 2014; Kühn et al 2022 |
| JPH3 | involved in regulating neuronal excitability and intracellular calcium signaling in the endoplasmic reticulum ; Double-knockout JPH3-4 mice show abnormal excitability and synaptic plasticity in hippocampal neurons ; Mutations implicated in Huntington disease–like 2 neurodegeneration | Garbino et al 2009; Kakizawa et al 2008; Bourinaris et al 2021 |
| PIGN | Involved in GPI-anchored protein synthesis; deficiencies in GPI anchors are associated with neurological abnormalities such as white matter degeneration; PIGN knockout mice show abnormal forebrain development, suggesting role in brain structure; PIGN mutations cause neurological phenotypes, including developmental delay, hypotonia, and epilepsy | Murakami et al 2025; McKean et al 2012; Ohba et al 2014 |
| REEP5 | knockdown of REEP5 together with RTN4 significantly reduces axonal ER tubules in cultured hippocampal neurons; mutationsin REEP5 cause hereditary spastic paraplegia; colocalizes with RTN3 immunoreactive dystrophic neurites in Alzheimer's brains | Yperman et al 2023; Sharoar et al 2019 |
| AMIGO3 | Associates to post-mortem brain tissue of AD ; inhibitory effect on oligodendrocyte precursor cell maturation, myelin production, and the axonal outgrowth of central nervous system neurons. | Jiang et al 2025; Foale et al 2017 |
| KIAA0513 | assumed to participate in neuroplasticity and apoptosis; expression is significantly downregulated in AD mouse models; | Lauriat et al 2006; Lian et al 2023 |
| XPO7 | Mice with *Xpo7* haploinsufficiency exhibit cognitive and social behavioral impairments; associates to risk of schizophrenia. | Toyoda et al 2025; Heinzer et al 2024 |
| TTI2 | Tti2 as a causal genetic link between glucose metabolism and structural brain plasticity; associates to neurodevelopmental disorders with microcephaly | Senko et al 2022; Serey-Gaut et al 2023 |
| MED30 | Part of the Mediator complex containing up to 30 proteins, is required for adult hippocampal neurogenesis and brain function in mice; parts of mediator complex are involved in various neurodegenerative diseases. | Chen et al 2020; Schiano et al 2023 |
| CC2D2B | Involved in cilium assembly; in brain cilium are involved in sensory detection and neuromodulation | Zhang et al 2012; Jurisch-Yaksi et al 2024 |
| SLX4IP | SLX4IP gene is activated by PAX7, which is a key transcription factor involved in neural development and oncogenesis in the nervous system; SLX4IP is involved in telomere maintenance; downregulated in AD mice when orally administrated phycocyanin. | Lee et al 2024; Robinson et al 2021; Imai et al 2021 |
| USP1 | Involved in morphogenesis of granule neuron dendrites and axons; Deubiquitinase. Regulating DNA repair processes | Anckar et al 2015; García-Santisteban et al 2013; |
| SCP2 | multipurpose lipid binding protein that shuttles cholesterol and other lipids from the endoplasmic reticulum; involved in endocannabinoid neurotransmitter system; regulates gamma-secretase which is involved in Alzheimer's disease pathology | Hillard et al 2017; Ko, M. H., & Puglielli, L. 2007 |
| RNPEP | Aminopeptidase B (RNPEP) processes glucagon and influences neuropeptide metabolism; involved in metabolic processes (proteomic changes) in AD context | Pham et al 2007; Mendonça et al 2019 |

| Gene | Association | Citation |
| --- | --- | --- |
| GCC2 | Golgin GCC185 (product of GCC2) is involved in endosomal lysosome enzyme recycling; Endolysosomal system may mediate AD risk. | Reddy et al 2006; Gao et al 2018 |
| CMSS1 | CMSS1 is modulator of immune system, which expression is increased in APOE4 male mice | Rao et al 2025 |
| MAK16 | Involved in astroglial development and axon pathfinding in zebrafish | Barresi et al 2010 |
| EMC7 | Endoplasmic reticulum membrane complex (EMC) supports GABAA receptor proteostasis (EMC7-containing core); hypothesis that EMC are involved in regulating various cellular processes, contributing to synaptic plasticity, neurodegeneration, axon regrowth, and information processing | Whittsette et al 2022; Khan et al 2022 |
| ZYG11A | an E3 ubiquitin ligase, a type of enzyme involved in protein degradation pathways that are often implicated in neurodegenerative disorders; associated with altered cytosine hydroxymethylation (CpH) in mice overexpressing SNCA, a key gene in Parkinson's disease; expression is altered in the hippocampus of rats engaged in voluntary wheel running, suggesting its involvement in brain function and aging-related processes | Schaffner et al 2023; Moon et al 2024 |
| ANAPC5 | APC/C complex subunit ANAPC5 participates in neuronal polarity and ciliary regulation; a transcription factor with reduced activity in affected AD samples | Ganner et al 2009; Sun et al 2019 |
| MEG9 | DLK1-DIO3 locus lncRNA cluster (including MEG9) is dynamically regulated by associative learning in mice; MEG9 is a lncRNA that was one of seven deregulated lncRNAs chosen for validation in post-mortem hippocampus samples from Alzheimer’s disease patients | Tan et al 2017; Filomena et al 2024 |
| JAG1 | (JAG1) is a Notch-ligand and its expression is repressed by FOXP1, a mechanism necessary for the neuronal differentiation of neural stem cells; astrocytes can negatively regulate nerve regeneration by influencing the Notch signaling pathway through JAG1; may be involved in the mechanisms of Alzheimer's Disease (AD) | Braccioli et al 2017; Magnusson et al 2014; Li et al 2022 |
| BCORL1 | a transcriptional co-repressor with a role in cortical migration, neuronal differentiation, maturation, and cerebellar development; one of the top five downregulated proteins in microglia from 5xFAD mice (an Alzheimer’s disease model) | Gafner et al 2022; Rangaraju et al 2018 |
| IRAG2/ LOC645177 | downregulated in an iPSC model of Jansen de Vries Syndrome (JdVS), a neurodevelopmental disorder; decreased levels in the Mild Cognitive Impairment-Dementia with Lewy Bodies (MCI-DLB) group; modulates the cAMP sensitivity of the HCN4 channel, which is highly expressed in the heart and brain – HCN channels play important roles in modulating cellular excitability, rhythmic activity, dendritic integration, and synaptic transmission | Aguilan et al 2023; Bellomo et al 2025; Peters et al 2020; Peters et al 2024; Ch |
| C4orf36 | associated with both schizophrenia (SCZ) and multiple sclerosis (MS) within a list of jointly associated loci; jointly associated with schizophrenia and vitamin D levels | Ahangari et al 2022; Jaholkowski et al 2023 |
| GP9 | upregulation of GP9, along with other structural subunits of the GPIb-IX-V platelet receptor in non-demented aged and individuals with AD; | de Sousa et al 2023 |

#### References to literature search for top periodic candidate gene set ($p < 0.00077$, n=32)

Aguilan, J. T., Pedrosa, E., Dolstra, H., Baykara, R. N., Barnes, J., Zhang, J., ... & Lachman, H. M. (2023). Proteomics and phosphoproteomics profiling in glutamatergic neurons and microglia in an iPSC model of Jansen de Vries Syndrome. bioRxiv.

Ahangari, M., Everest, E., Nguyen, T. H., Verrelli, B. C., Webb, B. T., Bacanu, S. A., ... & Riley, B. P. (2022). Genome-wide analysis of schizophrenia and multiple sclerosis identifies shared genomic loci with mixed direction of effects. Brain, Behavior, and Immunity, 104, 183-190.

Anckar, J., & Bonni, A. (2015). Regulation of neuronal morphogenesis and positioning by ubiquitin-specific proteases in the cerebellum. PLoS One, 10(1), e0117076.
Barresi, M. J., Burton, S., Dipietrantonio, K., Amsterdam, A., Hopkins, N., & Karlstrom, R. O. (2010). Essential genes for astroglial development and axon pathfinding during zebrafish embryogenesis. Developmental Dynamics, 239(10), 2603-2618.
Bellomo, G., Vermunt, L., In't Veld, S., Doecke, J., Hok-A-Hin, Y., Veverová, K., ... & del Campo, M. (2025). Plasma proteome profiling identified biomarkers for the differential diagnosis and molecular staging of neurodegenerative dementias.
Benitez, B. A., Jin, S. C., Guerreiro, R., Graham, R., Lord, J., Harold, D., ... & 3C Study Group. (2014). Missense variant in TREML2 protects against Alzheimer's disease. Neurobiology of aging, 35(6), 1510-e19.
Bourinaris, T., Athanasiou, A., Efthymiou, S., Wiethoff, S., Salpietro, V., & Houlden, H. (2021). Allelic and phenotypic heterogeneity in Junctophillin-3 related neurodevelopmental and movement disorders. European Journal of Human Genetics, 29(6), 1027-1031.
Braccioli, L., Vervoort, S. J., Adolfs, Y., Heijnen, C. J., Basak, O., Pasterkamp, R. J., ... & Coffer, P. J. (2017). FOXP1 promotes embryonic neural stem cell differentiation by repressing Jagged1 expression. Stem cell reports, 9(5), 1530-1545.
Chang, X., Wang, J., Jiang, H., Shi, L., & Xie, J. (2019). Hyperpolarization-activated cyclic nucleotide-gated channels: an emerging role in neurodegenerative diseases. Frontiers in molecular neuroscience, 12, 141.
Chen, G. Y., Zhang, S., Li, C. H., Qi, C. C., Wang, Y. Z., Chen, J. Y., ... & Su, C. J. (2020). Mediator Med23 regulates adult hippocampal neurogenesis. Frontiers in Cell and Developmental Biology, 8, 699.
Cheng, L., Liu, Z., Shen, C., Xiong, Y., Shin, S. Y., Hwang, Y., ... & Zhang, X. (2025). A Wonderful Journey: The Diverse Roles of Adenosine Deaminase Action on RNA 1 (ADAR 1) in Central Nervous System Diseases. CNS Neuroscience & Therapeutics, 31(1), e70208.
de Sousa, D. M. B., Poupardin, R., Villeda, S. A., Schroer, A. B., Fröhlich, T., Frey, V., ... & Kniewallner, K. M. (2023). The platelet transcriptome and proteome in Alzheimer’s disease and aging: an exploratory cross-sectional study. Frontiers in Molecular Biosciences, 10, 1196083.
Ding, Y. Q., Marklund, U., Yuan, W., Yin, J., Wegman, L., Ericson, J., ... & Chen, Z. F. (2003). Lmx1b is essential for the development of serotonergic neurons. Nature neuroscience, 6(9), 933-938.
Filomena, E., Picardi, E., Tullo, A., Pesole, G., & D’Erchia, A. M. (2024). Identification of deregulated lncRNAs in Alzheimer’s disease: an integrated gene co-expression network analysis of hippocampus and fusiform gyrus RNA-seq datasets. Frontiers in Aging Neuroscience, 16, 1437278.
Foale, S., Berry, M., Logan, A., Fulton, D., & Ahmed, Z. (2017). LINGO-1 and AMIGO3, potential therapeutic targets for neurological and dysmyelinating disorders?. Neural regeneration research, 12(8), 1247-1251.
Gafner, M., Michelson, M., Argilli, E., Yosovich, K., Sherr, E. H., Parks, K. C., ... & Blumkin, L. (2022). Major brain malformations: corpus callosum dysgenesis, agenesis of septum pellucidum and polymicrogyria in patients with BCORL1-related disorders. Journal of human genetics, 67(2), 95-101.
Ganner, A., Lienkamp, S., Schäfer, T., Romaker, D., Wegierski, T., Park, T. J., ... & Walz, G. (2009). Regulation of ciliary polarity by the APC/C. Proceedings of the National Academy of Sciences of the United States of America, 106(42), 17799.
Gao, S., Casey, A. E., Sargeant, T. J., & Mäkinen, V. P. (2018). Genetic variation within endolysosomal system is associated with late-onset Alzheimer’s disease. Brain, 141(9), 2711-2720.
Garbino, A., Van Oort, R. J., Dixit, S. S., Landstrom, A. P., Ackerman, M. J., & Wehrens, X. H. (2009). Molecular evolution of the junctophilin gene family. Physiological genomics, 37(3), 175-186.

García-Santisteban, I., Peters, G. J., Giovannetti, E., & Rodríguez, J. A. (2013). USP1 deubiquitinase: cellular functions, regulatory mechanisms and emerging potential as target in cancer therapy. Molecular cancer, 12(1), 91.
Hegde, A. N., Timm, L. E., Sivley, C. J., Ramiyaramcharankarthic, S., Lowrimore, O. J., Hendrix, B. J., ... & Anderson, W. J. (2025). Ubiquitin-Proteasome-Mediated Protein Degradation and Disorders of the Central Nervous System. International Journal of Molecular Sciences, 26(3), 966.
Heinzer, L., & Curtis, D. (2024). What have genetic studies of rare sequence variants taught us about the aetiology of schizophrenia?. Journal of Translational Genetics and Genomics, 8(1), 1-12.
Hillard, C. J., Huang, H., Vogt, C. D., Rodrigues, B. E., Neumann, T. S., Sem, D. S., ... & Cunningham, C. W. (2017). Endocannabinoid transport proteins: discovery of tools to study sterol carrier protein-2. In Methods in enzymology (Vol. 593, pp. 99-121). Academic Press.
Hu, Y. T., Chen, X. L., Huang, S. H., Zhu, Q. B., Yu, S. Y., Shen, Y., ... & Bao, A. M. (2019). Early growth response‐1 regulates acetylcholinesterase and its relation with the course of Alzheimer’s disease. Brain Pathology, 29(4), 502-512.
Imai, Y., Koseki, Y., Hirano, M., & Nakamura, S. (2021). Nutrigenomic studies on the ameliorative effect of enzyme-digested phycocyanin in alzheimer’s disease model mice. Nutrients, 13(12), 4431.
Jaholkowski, P., Hindley, G. F., Shadrin, A. A., Tesfaye, M., Bahrami, S., Nerhus, M., ... & Andreassen, O. A. (2023). Genome-wide Association Analysis of Schizophrenia and Vitamin D Levels Shows Shared Genetic Architecture and Identifies Novel Risk Loci. Schizophrenia Bulletin, 49(6), 1654-1664.
Jiang, W., Vogelgsang, J., Dan, S., Durning, P., McCoy, T. H., Berretta, S., & Klengel, T. (2025). Association of RDoC dimensions with post mortem brain transcriptional profiles in Alzheimer's disease. Alzheimer's & Dementia: Diagnosis, Assessment & Disease Monitoring, 17(2), e70103.
Jones, M. W., Errington, M. L., French, P. J., Fine, A., Bliss, T. V., Garel, S., ... & Davis, S. (2001). A requirement for the immediate early gene Zif268 in the expression of late LTP and long-term memories. Nature neuroscience, 4(3), 289-296.
Jurisch-Yaksi, N., Wachten, D., & Gopalakrishnan, J. (2024). The neuronal cilium–a highly diverse and dynamic organelle involved in sensory detection and neuromodulation. Trends in Neurosciences, 47(5), 383-394.
Kakizawa, S., Moriguchi, S., Ikeda, A., Iino, M., & Takeshima, H. (2008). Functional crosstalk between cell-surface and intracellular channels mediated by junctophilins essential for neuronal functions. The Cerebellum, 7(3), 385-391.
Kärkkäinen, V., Saari, T., Rusanen, M., Uusitalo, H., Leinonen, V., Thiede, B., ... & Utheim, T. P. (2025). Neuroinflammation Markers in Tear Fluid of Mild Alzheimer’s Disease. Journal of Molecular Neuroscience, 75(2), 73.
Khan, S. (2022). Endoplasmic reticulum in metaplasticity: from information processing to synaptic proteostasis. Molecular Neurobiology, 59(9), 5630-5655.
Ko, M. H., & Puglielli, L. (2007). The sterol carrier protein SCP-x/pro-SCP-2 gene has transcriptional activity and regulates the Alzheimer disease γ-secretase. Journal of Biological Chemistry, 282(27), 19742-19752.
Koemans, T. S., Kleefstra, T., Chubak, M. C., Stone, M. H., Reijnders, M. R., de Munnik, S., ... & Kramer, J. M. (2017). Functional convergence of histone methyltransferases EHMT1 and KMT2C involved in intellectual disability and autism spectrum disorder. PLoS genetics, 13(10), e1006864.
Kucukdereli, H., Allen, N. J., Lee, A. T., Feng, A., Ozlu, M. I., Conatser, L. M., ... & Eroglu, C. (2011). Control of excitatory CNS synaptogenesis by astrocyte-secreted proteins Hevin and SPARC. Proceedings of the National Academy of Sciences, 108(32), E440-E449.

Kühn, A. L., Frenzel, S., Teumer, A., Wittfeld, K., Garvert, L., Weihs, A., ... & Van der Auwera, S. (2022). TREML2 gene expression and its missense variant rs3747742 associate with white matter hyperintensity volume and Alzheimer's disease-related brain atrophy in the general population. International Journal of Molecular Sciences, 23(22), 13764.
Laguna, A., Schintu, N., Nobre, A., Alvarsson, A., Volakakis, N., Jacobsen, J. K., ... & Perlmann, T. (2015). Dopaminergic control of autophagic-lysosomal function implicates Lmx1b in Parkinson's disease. Nature Neuroscience, 18(6), 826-835.
Lauriat, T. L., Dracheva, S., Kremerskothen, J., Duning, K., Haroutunian, V., Buxbaum, J. D., ... & McInnes, L. A. (2006). Characterization of KIAA0513, a novel signaling molecule that interacts with modulators of neuroplasticity, apoptosis, and the cytoskeleton. Brain research, 1121(1), 1-11.
Lee, J., Kim, E., Kim, H., Kim, Y. J., & Kim, S. H. (2024). MEGF11 Activates RAD52‑Dependent ALT through the NELL2‑PAX7 Transcriptional Cascade during Malignant Transformation of MPNSTs.
Lee, K. S., Lee, T., Kim, M., Ignatova, E., Ban, H. J., Sung, M. K., ... & Choi, J. K. (2025). Shared rare genetic variants in multiplex autism families suggest a social memory gene under selection. Scientific Reports, 15(1), 696.
Li, F., Lin, Z., & Tian, G. (2022). Comprehensive analysis of lncRNA‑miRNA‑mRNA regulatory networks for Alzheimer's disease. Acta neurobiologiae experimentalis, 82(3), 263-272.
Lian, P., Cai, X., Wang, C., Liu, K., Yang, X., Wu, Y., ... & Xu, Y. (2023). Identification of metabolism-related subtypes and feature genes in Alzheimer's disease. Journal of Translational Medicine, 21(1), 628.
Madeo, M., Stewart, M., Sun, Y., Sahir, N., Wiethoff, S., Chandrasekar, I., ... & Kruer, M. C. (2016). Loss-of-function mutations in FRRS1L lead to an epileptic-dyskinetic encephalopathy. The American Journal of Human Genetics, 98(6), 1249-1255.
Magnusson, J. P., Göritz, C., Tatarishvili, J., Dias, D. O., Smith, E. M., Lindvall, O., ... & Frisén, J. (2014). A latent neurogenic program in astrocytes regulated by Notch signaling in the mouse. Science, 346(6206), 237-241.
McKean, D. M., & Niswander, L. (2012). Defects in GPI biosynthesis perturb Cripto signaling during forebrain development in two new mouse models of holoprosencephaly. Biology open, 1(9), 874-883.
Mendonça, C. F., Kuras, M., Nogueira, F. C. S., Plá, I., Hortobágyi, T., Csiba, L., ... & Rezeli, M. (2019). Proteomic signatures of brain regions affected by tau pathology in early and late stages of Alzheimer's disease. Neurobiology of disease, 130, 104509.
Moon, H. Y., & Lee, M. (2024). Exercise-induced expression of genes associated with aging in the hippocampus of rats. Neuroscience Letters, 823, 137646.
Murakami, Y. (2025). Biosynthesis of GPI anchored proteins, its deficiencies and treatment. Journal of Human Genetics, 1-9.
Ohba, C., Okamoto, N., Murakami, Y., Suzuki, Y., Tsurusaki, Y., Nakashima, M., ... & Saitsu, H. (2014). PIGN mutations cause congenital anomalies, developmental delay, hypotonia, epilepsy, and progressive cerebellar atrophy. neurogenetics, 15(2), 85-92.
Peters, C. H., Myers, M. E., Juchno, J., Haimbaugh, C., Bichraoui, H., Du, Y., ... & Proenza, C. (2020). Isoform-specific regulation of HCN4 channels by a family of endoplasmic reticulum proteins. Proceedings of the National Academy of Sciences, 117(30), 18079-18090.
Pham, V. L., Cadel, M. S., Gouzy-Darmon, C., Hanquez, C., Beinfeld, M. C., Nicolas, P., ... & Foulon, T. (2007). Aminopeptidase B, a glucagon-processing enzyme: site directed mutagenesis of the Zn2+-binding motif and molecular modelling. BMC biochemistry, 8(1), 21.
Rangaraju, S., Dammer, E. B., Raza, S. A., Gao, T., Xiao, H., Betarbet, R., ... & Seyfried, N. T. (2018). Quantitative proteomics of acutely-isolated mouse microglia identifies novel immune Alzheimer's disease-related proteins. Molecular neurodegeneration, 13(1), 34.

Rao, A., Chen, N., Kim, M. J., Blumenfeld, J., Yip, O., Liang, Z., ... & Huang, Y. (2025). Microglia depletion reduces human neuronal APOE4-related pathologies in a chimeric Alzheimer's disease model. Cell Stem Cell, 32(1), 86-104.
Reddy, J. V., Burguete, A. S., Sridevi, K., Ganley, I. G., Nottingham, R. M., & Pfeffer, S. R. (2006). A functional role for the GCC185 golgin in mannose 6-phosphate receptor recycling. Molecular biology of the cell, 17(10), 4353-4363.
Robinson, N. J., Miyagi, M., Scarborough, J. A., Scott, J. G., Taylor, D. J., & Schiemann, W. P. (2021). SLX4IP promotes RAP1 SUMOylation by PIAS1 to coordinate telomere maintenance through NF-κB and Notch signaling. Science signaling, 14(689), eabe9613.
Schaffner, S. L. (2023). Associations of DNA methylation with individual differences in Parkinson's disease susceptibility (Doctoral dissertation, University of British Columbia).
Schiano, C., Luongo, L., Maione, S., & Napoli, C. (2023). Mediator complex in neurological disease. Life Sciences, 329, 121986.
Senko, A. N., Overall, R. W., Silhavy, J., Mlejnek, P., Malínská, H., Hüttl, M., ... & Kempermann, G. (2022). Systems genetics in the rat HXB/BXH family identifies Tti2 as a pleiotropic quantitative trait gene for adult hippocampal neurogenesis and serum glucose. PLoS Genetics, 18(4), e1009638.
Serey-Gaut, M., Cortes, M., Makrythanasis, P., Suri, M., Taylor, A. M., Sullivan, J. A., ... & Antonarakis, S. E. (2023). Bi-allelic TTI1 variants cause an autosomal-recessive neurodevelopmental disorder with microcephaly. The American Journal of Human Genetics, 110(3), 499-515.
Sharoar, M. G., Hu, X., Ma, X. M., Zhu, X., & Yan, R. (2019). Sequential formation of different layers of dystrophic neurites in Alzheimer's brains. Molecular psychiatry, 24(9), 1369-1382.
Stewart, M., Lau, P., Banks, G., Bains, R. S., Castroflorio, E., Oliver, P. L., ... & Nolan, P. M. (2019). Loss of Frrs1l disrupts synaptic AMPA receptor function, and results in neurodevelopmental, motor, cognitive and electrographical abnormalities. Disease models & mechanisms, 12(2), dmm036806.
Sun, Q., Kong, W., Mou, X., & Wang, S. (2019). Transcriptional regulation analysis of Alzheimer's disease based on FastNCA algorithm. Current Bioinformatics, 14(8), 771-782.
Tan, M. C., Widagdo, J., Chau, Y. Q., Zhu, T., Wong, J. J. L., Cheung, A., & Anggono, V. (2017). The activity-induced long non-coding RNA Meg3 modulates AMPA receptor surface expression in primary cortical neurons. Frontiers in cellular neuroscience, 11, 124.
Toyoda, S., Kikuchi, M., Abe, Y., Tashiro, K., Handa, T., Katayama, S., ... & Shiwaku, H. (2025). Schizophrenia-related Xpo7 haploinsufficiency leads to behavioral and nuclear transport pathologies. EMBO reports, 26(4), 948-981.
Wang, S. Y., Fu, X. X., Duan, R., Wei, B., Cao, H. M., Chen, S. Y., ... & Jiang, T. (2023). The Alzheimer's disease-associated gene TREML2 modulates inflammation by regulating microglia polarization and NLRP3 inflammasome activation. Neural regeneration research, 18(2), 434-438.
Whittsette, A. L., Wang, Y. J., & Mu, T. W. (2022). The endoplasmic reticulum membrane complex promotes proteostasis of GABAA receptors. Iscience, 25(8).
Yperman, K., & Kuijpers, M. (2023). Neuronal endoplasmic reticulum architecture and roles in axonal physiology. Molecular and Cellular Neuroscience, 125, 103822.
Zhang, D., & Aravind, L. (2012). Novel transglutaminase-like peptidase and C2 domains elucidate the structure, biogenesis and evolution of the ciliary compartment. Cell cycle, 11(20), 3861-3875.
Peters, C. H., Singh, R. K., Langley, A. A., Nichols, W. G., Ferris, H. R., Jeffrey, D. A., ... & Bankston, J. R. (2024). LRMP inhibits cAMP potentiation of HCN4 channels by disrupting intramolecular signal transduction. Elife, 12, RP92411.

### Supplementary Table S2 Literature search for top aperiodic candidate gene set ($p < 0.00077$, n=20)

| Gene | Association | Citation |
|---|---|---|
| ALDH1A2 | Decrease of ALDH1A2 expression in mice is associated to neuron death; ALDH gene superfamily enzymes play a role in modulating cell proliferation, differentiation, and survival | Liang, H. et al. 2017; Grünblatt, E. 2016 |
| APOBEC3A | RNA C-to-U editing (sites) by APOBECs is associated with neurological diseases such as Alzheimer's disease and epilepsy; | Min, D. J. et al 2026; Van Norden, M et al 2023 |
| BCL7B | Possible role for BCL7B in oligodendrogenesis; maps to exercise-responsive ageing pathways in humans | Wischhof, L.et al 2022; Juan, C. G. et al 2026 |
| C5AR2 | Part of complement system that has roles in neurodevelopment, adult neural plasticity, neuroprotection and neuroinflammation. C5AR2 involved in neuroprotection. Expressed in microglia and possible role in microglial activation. | Pekna, M. et al 2021; Bohlson, S. S. et al 2023; Nguyen, V. D. et al 2025 |
| CALN1 | Role in neurodevelopment. Mice with CALN1 knockout leads to gene dysregulation related to forebrain development. Hub gene involved in AD, may regulate trans-synaptic signalling. upregulated in the hippocampus of stressed/depressed mice. | Li, H. J. et al 2024; Huang, Z. et al 2022; Pavlova, M. B. et al 2023 |
| EEF1A1 | Reduction of EEF1A1 in excitatory synapses in mice overexpressin human alpha-synuclein; reduced expression in AD brain; critical for maintaining long-term synaptic plasticity, cellular model for learning and memory | Blumenstock, S. et al 2019; Beckelman, B. C. et al 2016 |
| EMB | EMB has a suggested role in neuroinflammation; potential biomarker for Alzheimer's disease | Agrawal, S. M., & Yong, V. W. 2011; Zheng, D. et al 2022 |
| IQSEC1 | Hippocampal gene critical for maintaining excitatory synapses through AMPA and NMDA receptor trafficking and regulating synaptic long-term depression. | Brabec, J. L. et al 2021 |
| MFSD14A | MFSD14A gene expression is downregulated in the hypothalamus and brainstem of mice subjected to food starvation; Differentially expressed mRNAs detected in exosomes from Alzheimer's disease mice | Lekholm, E. et al 2017; Su, L. et al 2022 |
| NCF1 | NCF1 deficiency attenuates TBI-induced inflammatory responses in knockout mice; differentially expressed genes in the striatum of Parkinson's disease mouse model; May interact with and regulate the activity of the E3 ubiquitin ligase ZNRF1, which promotes neurite degeneration | Gao, T. X. et al 2024; Ishaq, S. et al 2025; Wakatsuki, S., & Araki, T. 2023 |
| NFYB | As a part of NF-Y complex, essential for neural progenitor maintenance during brain morphogenesis. NF-Y is implicated in neurodegeneration | Yamanaka, T. et al 2024 |
| OMD | Associated to Parkinson's disease, a common neurodegenerative disease; Altered RNA transcription in white matter lesions linked to neurodegeneration. | Surguchov, A. 2021; Simpson, J. E. et al 2009 |
| PHLDA1 | Role in neurological diseases. Promotes neuroinflammation, mitochondrial dysfunction, endoplasmic reticulum stress in neurons. | Liu, X. et al 2026 |
| RRAGD | Inhibition of mTOR signalling is associated with overexpression of RRAGD. mTOR signaling cascade plays a key role in the development, synaptic plasticity, memory, and metabolic regulation of the central nervous system (CNS); mTOR signalling pathway is implicated in neurodegeneration. | Movahedpour, A. et al 2022; Querfurth, H., & Lee, H. K. 2021 |
| SCRG1 | SCRG1 is a marker for astrocyte subtype which accumulate autophagosomes and appear in the aging hippocampus of mice; Implicated role in transmissible Spongiform Encephalopathies, neurodegenerative diseases. | Lee, E. et al 2022; Sahu, P. S., & Ter, E. 2018 |
| SF3B2 | Methylation of SF3B2 is essential for synapse development, learning, and memory in a mouse model. Reducing SF3B2 expression in neuronal cells is neuroprotective. | Hashimoto, M. 2026; Zhu, J. et al 2016 |
| TMEM271 | Differentially downregulated in dementia. Expressed in brain. | Vastrad, B., & Vastrad, C. 2025; Uhlén, M. et al 2015 |
| TREML2 | Modulates inflammation by regulating microglial polarization and NLRP3 inflammasome activation; Protective TREML2 missense variant associates with reduced Alzheimer's disease risk; associates with whitematter hyperintensity volume | Wang et al 2023; Benitez et al 2014; Kühn et al 2022 |
| XKR6 | XKR6 locus associated with White Matter Hyperintensities. Risk gene related to abnormal cortical structural indicators in patients with subcortical ischemic vascular disease and cognitive impairment. | Sargurupremraj, M. et al 2020; Huang, J. et al 2025 |
| ZFP37 | Zebrafish ZNF32 homolog suggested to be involved in nerve regeneration; belongs to KZNF genes that are involved in neurodevelopment. | Wei, Y. et al 2016; Farmiloe, G. et al 2020 |

**References for literature search of the top aperiodic candidate gene set ($p < 0.00077$, n=20)**


Agrawal, S. M., & Yong, V. W. (2011). The many faces of EMMPRIN—roles in neuroinflammation. Biochimica et Biophysica Acta (BBA)-Molecular Basis of Disease, 1812(2), 213-219.

Beckelman, B. C., Zhou, X., Keene, C. D., & Ma, T. (2016). Impaired eukaryotic elongation factor 1A expression in Alzheimer's disease. Neurodegenerative Diseases, 16(1-2), 39-43.

Benitez, B. A., Jin, S. C., Guerreiro, R., Graham, R., Lord, J., Harold, D., ... & 3C Study Group. (2014). Missense variant in TREML2 protects against Alzheimer's disease. Neurobiology of aging, 35(6), 1510-e19.

Blumenstock, S., Angelo, M. F., Peters, F., Dorostkar, M. M., Ruf, V. C., Luckner, M., ... & Herms, J. (2019). Early defects in translation elongation factor 1α levels at excitatory synapses in α-synucleinopathy. Acta neuropathologica, 138(6), 971-986.

Bohlson, S. S., & Tenner, A. J. (2023). Complement in the brain: contributions to neuroprotection, neuronal plasticity, and neuroinflammation. Annual Review of Immunology, 41(1), 431-452.

Brabec, J. L., Lara, M. K., Tyler, A. L., & Mahoney, J. M. (2021). System-level analysis of Alzheimer’s disease prioritizes candidate genes for neurodegeneration. Frontiers in Genetics, 12, 625246.

Farmiloe, G., Lodewijk, G. A., Robben, S. F., van Bree, E. J., & Jacobs, F. M. (2020). Widespread correlation of KRAB zinc finger protein binding with brain-developmental gene expression patterns. Philosophical Transactions of the Royal Society B: Biological Sciences, 375(1795).

Gao, T. X., Liang, Y., Li, J., Zhao, D., Dong, B. J., Xu, C., ... & Zhao, C. S. (2024). Knockout of neutrophil cytosolic factor 1 ameliorates neuroinflammation and motor deficit after traumatic brain injury. Experimental Neurology, 382, 114983.

Grünblatt, E., & Riederer, P. (2016). Aldehyde dehydrogenase (ALDH) in Alzheimer’s and Parkinson’s disease. Journal of Neural Transmission, 123(2), 83-90.

Hashimoto, M. (2026). Protein Arginine Methylation in the Nervous System Development, Health and Disease. In Biotechnological Advances in Healthomics (pp. 135-157). Singapore: Springer Nature Singapore.

Huang, J., Cheng, R., Liu, X., Chen, L., & Luo, T. (2025). Association of cortical macrostructural and microstructural changes with cognitive performance and gene expression in subcortical ischemic vascular disease patients with cognitive impairment. Brain Research Bulletin, 222, 111239.

Huang, Z. H., Wang, H., Wang, D. M., Zhao, X. Y., Liu, W. W., Zhong, X., ... & Lu, M. H. (2022). Identification of core genes in prefrontal cortex and hippocampus of Alzheimer's disease based on mRNA‐miRNA network. Journal of Cellular and Molecular Medicine, 26(23), 5779-5793.

Ishaq, S., Shah, I. A., Lee, S. D., & Wu, B. T. (2025). Transcriptomic Analysis of Immune-Related Genes in the Striatum of Parkinson’s Disease Brain Across Mouse Strains. Journal of Molecular Neuroscience, 75(3), 96.

Juan, C. G., & Ntasis, L. (2026). Multi-omic deep learning identifies exercise-responsive ageing pathways in humans. medRxiv, 2025-12.

Kühn, A. L., Frenzel, S., Teumer, A., Wittfeld, K., Garvert, L., Weihs, A., ... & Van der Auwera, S. (2022). TREML2 gene expression and its missense variant rs3747742 associate with white matter hyperintensity volume and Alzheimer’s disease-related brain atrophy in the general population. International Journal of Molecular Sciences, 23(22), 13764.

Lee, E., Jung, Y. J., Park, Y. R., Lim, S., Choi, Y. J., Lee, S. Y., ... & Chung, W. S. (2022). A distinct astrocyte subtype in the aging mouse brain characterized by impaired protein homeostasis. Nature Aging, 2(8), 726-741.

Lekholm, E., Perland, E., Eriksson, M. M., Hellsten, S. V., Lindberg, F. A., Rostami, J., & Fredriksson, R. (2017). Putative membrane-bound transporters MFSD14A and MFSD14B are neuronal and affected by nutrient availability. Frontiers in Molecular Neuroscience, 10, 11.
Li, H. J., Yu, X., Liu, X., Xu, J., Chen, J., Cheng, T., ... & Shao, Z. (2024). Calneuron 1 reveals the pivotal roles in schizophrenia via perturbing human forebrain development and causing hallucination-like behavior in mice. bioRxiv, 2024-04.
Liang, H., Wu, C., Deng, Y., Zhu, L., Zhang, J., Gan, W., ... & Xu, R. (2017). Aldehyde dehydrogenases 1A2 expression and distribution are potentially associated with neuron death in spinal cord of Tg (SOD1* G93A) 1Gur mice. International Journal of Biological Sciences, 13(5), 574.
Liu, X., Lv, Z., Xu, G., Chen, Y., Liu, H., & Xu, P. (2026). Investigating the Multiple Regulatory Mechanisms and Therapeutic Targets of PHLDA1 in Neurological Diseases. Current Neuropharmacology.
Min, D. J., Lee, S., Lee, Y. S., & Cho, J. (2026). Two codes of RNA editing by deamination in human diseases. Experimental & Molecular Medicine, 1-14.
Movahedpour, A., Vakili, O., Khalifeh, M., Mousavi, P., Mahmoodzadeh, A., Taheri‐Anganeh, M., ... & Khatami, S. H. (2022). Mammalian target of rapamycin (mTOR) signaling pathway and traumatic brain injury: A novel insight into targeted therapy. Cell biochemistry and function, 40(3), 232-247.
Nguyen, V. D., Bright, M., Zhou, Y., Morgan, B. P., & Zelek, W. M. (2025). Complement biosynthesis in human brain: Insights from single-nucleus transcriptomics of hippocampus. BioRxiv, 2025-05.
Pavlova, M. B., Smagin, D. A., Kudryavtseva, N. N., & Dyuzhikova, N. A. (2023). Changes in Expression of Genes Associated with Calcium Processes in the Hippocampus in Mice Exposed to Chronic Social Stress. Molecular Biology, 57(2), 356-365.
Pekna, M., & Pekny, M. (2021). The complement system: a powerful modulator and effector of astrocyte function in the healthy and diseased central nervous system. Cells, 10(7), 1812.
Querfurth, H., & Lee, H. K. (2021). Mammalian/mechanistic target of rapamycin (mTOR) complexes in neurodegeneration. Molecular neurodegeneration, 16(1), 44.
Sahu, P. S., & Ter, E. (2018). Interactions between neurotropic pathogens, neuroinflammatory pathways, and autophagic neural cell death. Neuroimmunology and Neuroinflammation, 5, N-A.
Sargurupremraj, M., Suzuki, H., Jian, X., Sarnowski, C., Evans, T. E., Bis, J. C., ... & Debette, S. (2020). Cerebral small vessel disease genomics and its implications across the lifespan. Nature communications, 11(1), 6285.
Simpson, J. E., Hosny, O., Wharton, S. B., Heath, P. R., Holden, H., Fernando, M. S., ... & Ince, P. G. (2009). Microarray RNA expression analysis of cerebral white matter lesions reveals changes in multiple functional pathways. Stroke, 40(2), 369-375.
Su, L., Li, R., Zhang, Z., Liu, J., Du, J., & Wei, H. (2022). Identification of altered exosomal microRNAs and mRNAs in Alzheimer's disease. Ageing research reviews, 73, 101497.
Surguchov, A. (2021). Biomarkers in Parkinson’s disease. In Neurodegenerative diseases biomarkers: Towards translating research to clinical practice (pp. 155-180). New York, NY: Springer US.
Uhlén, M., Fagerberg, L., Hallström, B. M., Lindskog, C., Oksvold, P., Mardinoglu, A., ... & Pontén, F. (2015). Tissue-based map of the human proteome. Science, 347(6220), 1260419. https://www.proteinatlas.org/ENSG00000273238-TMEM271. Version 25.0

Van Norden, M., Falls, Z., Mandloi, S., Segal, B., Baysal, B., Samudrala, R., & Elkin, P. L. (2023). The Role of C-to-U RNA Editing in Human Biodiversity. bioRxiv.
Vastrad, B., & Vastrad, C. (2025). Identification of key pathways and genes in dementia via integrated Bioinformatics analysis. SN Comprehensive Clinical Medicine, 7(1), 237.
Wakatsuki, S., & Araki, T. (2023). Novel insights into the mechanism of reactive oxygen species-mediated neurodegeneration. Neural Regeneration Research, 18(4), 746-749.
Wang, S. Y., Fu, X. X., Duan, R., Wei, B., Cao, H. M., Chen, S. Y., ... & Jiang, T. (2023). The Alzheimer's disease-associated gene TREML2 modulates inflammation by regulating microglia polarization and NLRP3 inflammasome activation. Neural regeneration research, 18(2), 434-438.
Wei, Y., Li, K., Yao, S., Gao, J., Li, J., Shang, Y., ... & Wei, Y. (2016). Loss of ZNF32 augments the regeneration of nervous lateral line system through negative regulation of SOX2 transcription. Oncotarget, 7(43), 70420.
Wischhof, L., Lee, H. M., Tutas, J., Overkott, C., Tedt, E., Stork, M., ... & Bano, D. (2022). BCL7A‐containing SWI/SNF/BAF complexes modulate mitochondrial bioenergetics during neural progenitor differentiation. The EMBO journal, 41(23), EMBJ2022110595.
Yamanaka, T., Kurosawa, M., Yoshida, A., Shimogori, T., Hiyama, A., Maity, S. N., ... & Nukina, N. (2024). The transcription factor NF-YA is crucial for neural progenitor maintenance during brain development. Journal of Biological Chemistry, 300(2).
Zheng, D., Tahir, R. A., Yan, Y., Zhao, J., Quan, Z., Kang, G., ... & Qing, H. (2022). Screening of human circular RNAs as biomarkers for early onset detection of Alzheimer's disease. Frontiers in Neuroscience, 16, 878287.
Zhu, J., Carozzi, V. A., Reed, N., Mi, R., Marmiroli, P., Cavaletti, G., & Hoke, A. (2016). Ethoxyquin provides neuroprotection against cisplatin-induced neurotoxicity. Scientific reports, 6(1), 28861.

**Supplementary Table S3 Descriptive statistics of AD associated genes and variants**

| GENE | RSID | Functional Consequence | CHR | POS | A1 | A2 | HomAlt_Count | Het_Count | HomRef_Count | MAF |
|---|---|---|---|---|---|---|---|---|---|---|
| C5AR2 | rs36046934 | synonymous_variant | 19 | 47341396 | A | G | 0 | 3 | 189 | 0.008 |
| RASGEF1C | rs11546322 | synonymous_variant | 5 | 180136435 | C | T | 18 | 93 | 81 | 0.336 |
| RASGEF1C | rs149114740 | synonymous_variant | 5 | 180118808 | T | C | 0 | 2 | 190 | 0.005 |
| RASGEF1C | rs7725201 | stop_gained | 5 | 180102124 | G | A | 14 | 89 | 89 | 0.305 |
| TREML2 | rs115991880 | missense_variant | 6 | 41194466 | T | G | 0 | 4 | 188 | 0.010 |
| TREML2 | rs3747742 | missense_variant | 6 | 41194780 | C | T | 21 | 79 | 92 | 0.315 |
| TREML2 | rs41273768 | missense_variant | 6 | 41198161 | A | G | 0 | 14 | 178 | 0.036 |
| TREML2 | rs41273770 | synonymous_variant | 6 | 41198239 | G | A | 0 | 11 | 181 | 0.029 |
| TREML2 | rs41273772 | missense_variant | 6 | 41198417 | C | T | 3 | 41 | 148 | 0.122 |
| TREML2 | rs61734887 | missense_variant | 6 | 41194824 | C | G | 0 | 14 | 178 | 0.036 |
| TREML2 | rs61998254 | synonymous_variant | 6 | 41198413 | G | A | 3 | 41 | 148 | 0.122 |
| TREML2 | rs62396355 | missense_variant | 6 | 41198411 | G | A | 3 | 41 | 148 | 0.122 |
| TREML2 | rs77704965 | missense_variant | 6 | 41198416 | C | G | 3 | 41 | 148 | 0.122 |

#### Supplementary Table S4 Top (P < 0.00077) Periodic And Aperiodic Gene Characterization

| Category | Periodic (n=32) | Aperiodic (n=20) |
|---|---|---|
| **1. Synaptic Transmission & Excitability** | 9 (28.1%) | 5 (25.0%) |
| **2. Neuronal Differentiation & Structure** | 12 (37.5%) | 7 (35.0%) |
| **3. Metabolism, ER Stress & Homeostasis** | **11 (34.4%)** | 3 (15.0%) |
| **4. Neuroinflammation & Immune Regulation** | 6 (18.8%) | **6 (30.0%)** |
| **5. Gene Regulation & Epigenetics** | 7 (21.9%) | 4 (20.0%) |
| **6. Vascular / Other** | 4 (12.5%) | 3 (15.0%) |

#### Detailed Gene Classification

##### 1. Synaptic Transmission & Excitability

Periodic (9): SPARCL1 (Excitatory synaptogenesis), FRRS1L (AMPA assembly), EGR1 (LTP/Memory), ADAR (Synaptic plasticity/Homeostasis), JPH3 (Excitability/Intracellular Ca2+), KIAA0513 (Neuroplasticity), SCP2 (Endocannabinoid system), EMC7 (GABA-A proteostasis), IRAG2 (HCN4 channels/Excitability).

Aperiodic (5): CALN1 (Trans-synaptic signalling), EEF1A1 (Excitatory synapses/Plasticity), IQSEC1 (AMPA & NMDA trafficking/LTD), RRAGD (Synaptic plasticity/mTOR), SF3B2 (Synapse development/Learning).

##### 2. Neuronal Differentiation & Structure

Periodic (12): KMT2C (Learning/Memory development), PIGN (Forebrain/White matter development), REEP5 (Axonal ER tubules), AMIGO3 (Axonal outgrowth/Myelin inhibition), TTI2 (Structural brain plasticity), MED30 (Hippocampal neurogenesis), CC2D2B (Brain cilium assembly), USP1 (Dendrite & Axon morphogenesis), MAK16 (Axon pathfinding), ANAPC5 (Neuronal polarity/Ciliary), JAG1 (Neural stem cell differentiation), BCORL1 (Cortical migration/Maturation).

Aperiodic (7): BCL7B (Oligodendrogenesis), C5AR2 (Neurodevelopment/Adult plasticity), CALN1 (Forebrain development), NFYB (Neural progenitor maintenance), OMD (White matter lesions), XKR6 (Cortical structure/White matter), ZFP37 (Nerve regeneration).

##### 3. Metabolism, ER Stress & Homeostasis

Periodic (11): LMX1B (Autophagic-lysosomal pathway), ADAR (Neuronal homeostasis), JPH3 (ER calcium signalling), REEP5 (Axonal ER network), TTI2 (Glucose metabolism), USP1 (Ubiquitin-proteasome pathway), SCP2 (Cholesterol/Lipid shuttling), RNPEP (Neuropeptide processing), GCC2 (Endolysosomal recycling), EMC7 (ER membrane complex/Proteostasis), ZYG11A (Protein degradation/E3 ligase).

Aperiodic (3): PHLDA1 (Mitochondrial dysfunction/ER stress), RRAGD (CNS metabolic regulation/mTOR), SCRG1 (Autophagosome accumulation).

##### 4. Neuroinflammation & Immune Regulation

Periodic (6): SPARCL1 (Astrocyte-secreted factors), ADAR (Innate immune regulation), TREML2 (Microglial polarization/Inflammasome), CMSS1 (Immune system modulator), JAG1 (Astrocyte-regulated nerve regeneration), BCORL1 (Microglia transcription).

Aperiodic (6): C5AR2 (Microglial activation/Complement), EMB (Neuroinflammation), NCF1 (TBI-induced inflammation), PHLDA1 (Neuroinflammation promoter), SCRG1 (Aging astrocyte marker), TREML2 (Microglial polarization).

##### 5. Gene Regulation & Epigenetics

Periodic (7): KMT2C (H3K4 methyltransferase), ADAR (RNA editing), SLX4IP (PAX7-activated transcription), ZYG11A (Cytosine hydroxymethylation), ANAPC5 (Transcription factor), MEG9 (Dynamically regulated lncRNA), BCORL1 (Transcriptional co-repressor).

Aperiodic (4): APOBEC3A (RNA C-to-U editing), EMB (Neuroprotective Circular RNA), OMD (Altered RNA transcription), SF3B2 (Methylation for memory).

**6. Vascular / Other**

Periodic (4): FRRS1L (Epilepsy/Autism target), XPO7 (Schizophrenia risk), C4orf36 (Schizophrenia/Vitamin D), GP9 (Platelet receptor/AD brain).

Aperiodic (3): APOBEC3A (Epilepsy), MFSD14A (Hypothalamic starvation response), XKR6 (Ischemic vascular disease/White matter).

#### Supplementary Table S5 Between-cluster comparison of key features after k-means clustering on aperiodic gene variants.

| Feature | F Statistic | P Value | P Adjusted |
|---|---|---|---|
| age | 2580.5 | 0.206 | 0.918 |
| sex | 2808 | 0.554 | 0.918 |
| education | 3246.5 | 0.324 | 0.918 |
| MOCA | 3223 | 0.382 | 0.918 |
| RAVLT | 2992.5 | 0.918 | 0.918 |
| BNT | 3063 | 0.512 | 0.918 |
| CFLUENCY | 3651 | 0.022 | 0.219 |
| TMTB | 2850 | 0.714 | 0.918 |
| APOE_risk | 3046 | 0.754 | 0.918 |
| p-tau217 | 2909 | 0.864 | 0.918 |

P values are based on the Wilcoxon test; Benjamini-Hochberg was used to correct for multiple tests. Abbreviations: APOE_risk = risk of developing dementia based on APOE allele (range 1–5). MOCA = Montreal cognitive assessment. RAVLT = Rey auditory verbal learning test - delayed recall. BNT = Boston Naming Test. CFLUENCY = Category Fluency Test. TMTB = Trail Making Test Part B. p-tau217 = phosphorylated Tau 217 plasma concentration.

**Supplementary Table S6 Top periodic candidate gene set (p < 0.005, n=145)**

| | | | | | |
|---|---|---|---|---|---|
| ACAN | CMSS1 | HDAC5 | MED30 | RASAL2 | TMEM45B |
| ACY3 | COL5A2 | HEYL | MEG9 | RASGEF1C | TPX2 |
| ADAR | CPSF1 | HHIPL2 | MIER1 | REEP5 | TREML2 |
| AFMID | CWC27 | HP | MKRN1 | REN | TRIP6 |
| AKT1S1 | CXCL16 | HTRA4 | MROH1 | RNPEP | TTI2 |
| AMIGO3 | CYP2B6 | IGF2BP3 | MRPL20 | RRM2B | TWSG1 |
| ANAPC5 | DNAI1 | INPP5B | MSR1 | RSBN1L | TYMSOS |
| ANTXR1 | EGR1 | INSL5 | MTPN | RSPO3 | UNC45B |
| ARHGAP44 | EMC7 | JAG1 | NOL4 | SCP2 | UQCRHL |
| BCORL1 | FAIM | JPH3 | NPAS2 | SEC22B | USP1 |
| C4orf36 | FAM169B | KBTBD3 | OR5AC2 | SERPINB9 | USP40 |
| CA10 | FFAR4 | KCNQ1 | OR7A10 | SH3PXD2A | VENTX |
| CABIN1 | FGF4 | KIAA0513 | OSCAR | SLC15A2 | VSIG10 |
| CAMKK2 | FLT3 | KIF1C | PDCD2 | SLC27A4 | XPO7 |
| CAVIN1 | FRRS1L | KLF8 | PI4KA | SLFN12L | ZBTB47 |
| CC2D2B | FUT8 | KMT2C | PIGN | SLX4IP | ZNF486 |
| CCDC169 | GABRA1 | LIG3 | POFUT2 | SPARCL1 | ZNF543 |
| CCKAR | GALNT4 | LMNB1 | PPP1R2 | SPATA13 | ZNF551 |
| CDC20B | GCC2 | LMX1B | PPWD1 | SPATA48 | ZNF71 |
| CDCP1 | GGTA1 | LOC645177 | PRR20G | SPIDR | ZYG11A |
| CDH6 | GMEB2 | MACROD1 | PRR23C | TAC4 | |
| CEP295 | GP2 | MAK16 | PRSS50 | TACSTD2 | |
| CER1 | GP9 | MAN2A2 | PUSL1 | TEDC1 | |
| CHURC1-FNT | GRIFIN | MBNL2 | RAB7B | TMC4 | |
| CLC | GSN | | RAD9B | TMEM229B | |
| | GTF2F2 | | | | |

**Supplementary Table S7 Top aperiodic candidate gene set ($p < 0.005$, n=39)**

| | |
|---|---|
| ACBD7-DCLRE1CP1 | OMD |
| ALDH1A2 | OTUD7B |
| APOBEC3A | PCNA |
| BCL7B | PCYT1B |
| C4orf51 | PHLDA1 |
| C5AR2 | RALGPS2 |
| CALN1 | RGR |
| COQ4 | RNASE8 |
| EEF1A1 | RRAGD |
| EMB | SCRG1 |
| FCRL6 | SEZ6 |
| INSM2 | SF3B2 |
| IQSEC1 | TDRD10 |
| LRRC10 | TMEM271 |
| MAP1LC3B | TREML2 |
| MFSD14A | UPK3B |
| NCF1 | XKR6 |
| NFXL1 | ZFP37 |
| NFYB | ZNF556 |
| NOL4 | |

#### Literature review of the candidate genes

RASGEF1C belongs to a family of RASGEF1 genes that encode proteins involved in Ras protein signal transduction, a pathway involved in synaptic plasticity in neuronal cells.[1,2] RASGEF1C is expressed in the brain,(Fagerberg et al., 2014) and for some transcripts, the functional variant rs7725201 in our study results in a stop codon (Supplementary Table 3). This variant is located in the Ras-GEF domain, which is vital for activating the Ras signalling pathway. TREML2 belongs to the triggering receptors expressed on the myeloid cells (TREM) family, along with TREM1 and TREM2.[4] In contrast to the plasma anti-inflammatory effects of TREM1 and TREM2, TREML2 seems to have a pro-inflammatory response.[5] Interestingly, the rs3747742 variant, included in our functional variants of TREML2, is associated with white matter hyperintensity and with the protective effect of AD.[6] Complement component 5a receptor C5AR2 is upregulated in mice model of AD and neuroprotective in mice model of spinal cord injury.[7,8] It also downregulates C5AR1 which has been suggested as a therapeutic target for AD.[9,10]

Out of 32 top candidate genes ($P < 0.00077$) associated with the periodic EEG features, 9 (28.1%) had implications in synaptic transmission and neuronal excitability, and 12 (37.5%) in neurodevelopmental processes (neuronal differentiation and structure). Similarly, out of 20 top aperiodic candidate genes ($P < 0.00077$), the counts were 5 (25%) and 7 (35%), respectively. These include FRRS1L, which is required for the assembly of the glutamatergic neurotransmission receptor AMPA.[11] Interestingly, Frrs1l knockout mice had abnormal EEG showing repeated runs of polyspikes and seizure-like episodes. SPARCL1 is secreted by astrocytes and induces synapse formation.[12] Sparcl1 knockout mice have fewer excitatory synapses than wild-type mice. Mice deficient in Erg1 had impaired late long-term potentiation in the dentate gyrus, although early long-term potentiation was present.[13] Therefore, ERG1 is essential for the transition from short- to long-term synaptic plasticity and the expression of long-term memories. In the prefrontal cortex of postmortem human samples, the expression of ERG1-mRNA together with AChE-mRNA seems to decrease in late AD stages compared to early AD, and the study suggests that EGR1 can upregulate AChE expression.[14] JPH3 is involved in the regulation of neuronal excitability and intracellular calcium signalling pathways.[15] Double-knockout JPH3-4 mice show abnormal excitability and synaptic plasticity in hippocampal neurons.[16] IRAG2 modulates the cAMP sensitivity of the HCN4 channels.[17] HCN4 channels play important roles in modulating cellular excitability, rhythmic activity, dendritic integration and synaptic transmission.[18] EMC7 is a subunit of the endoplasmic reticulum membrane complex involved in the biogenesis and function of GABA-A receptors, the primary inhibitory ion channels in the mammalian brain.[19] MEG9 is part of an lncRNA cluster and is expressed in response to glycine stimulation in a manner dependent on N-methyl-D-aspartate glutamate receptors (NMDA).[20] MEG9 expression levels are elevated during associative learning in mice. Furthermore, in the hippocampus of postmortem AD patients MEG9 is deregulated.[21] KIAA0513 interacts with proteins involved in neuroplasticity, apoptosis, and the cytoskeleton and is therefore suggested to be involved in signalling pathways related to these processes.[22] The KIAA0513 gene is expressed at low levels in AD and correlates with its progression.[23] ADAR is involved in RNA editing, neuronal homeostasis, synaptic plasticity, and innate immune regulation, and its dysfunction is associated with several central nervous system diseases, including AD.[24]

When comparing the top aperiodic to top periodic candidate genes, the periodic candidate genes are more heavily involved ( 34.4% vs 15% ) in the intracellular maintenance of neurons. SCP2 is a lipid binding protein that functions as a regulator in the endocannabinoid neurotransmitter system.[25] Interestingly, SCP2 also regulates the transcription of CD147, the regulatory subunit of the γ-secretase involved in amyloid β-peptide formation in

Alzheimer’s disease.[26] JPH3 on the other hand is a junctophilin encoding gene expressed highly in the hippocampus and involved in ER calcium signaling.[15] JPH3 is also associated with Huntington's disease. LMX1B is a transcription factor required for the normal execution of the autophagic-lysosomal pathway and also required for the development of serotonergic neurons.[27,28] USP1 is a deubiquitinase, involved in morphogenesis of granule neuron dendrites and axons and regulating DNA repair processes.[29,30] ZYG11A is an E3 ubiquitin ligase that is associated with altered methylation in Parkinson’s disease mice model.[31] Interestingly, its expression is altered in the hippocampus of rats engaged in exercise. [32] GCC2 is involved in mannose 6-phosphate receptor (MPR) recycling from endosomes to trans-Golgi network.[33] MPRs deliver lysosomal enzymes to endosomes and are therefore a vital part of endolysosomal system. Genetic variation in endolysosomal system has been associated with AD.[34]

Aperiodic candidate genes on the other hand are more involved in neuroinflammation ( 30% vs 18.8% ). C5AR2 is a part of the complement system encoding receptor for C5a neuroinflammatory chemoattractant and in the hippocampus it is predominantly expressed in microglia.[35,36] Interestingly, C5AR2 has suggested a neuroprotective in AD model and its expression is also decreased in AD. EMB is a member of the immunoglobulin superfamily and has a suggested role in neuroinflammation.[37] Interestingly, EMB is also involved in neurodevelopment and long-term memory formation and interacts with a circular-RNA suggested as a biomarker for AD.[38] Knockout of neutrophil cytosolic factor 1, NCF1, protects traumatic brain injury induced mice from neuroinflammation.[39] Interestingly, NCF1 also initiates ZNRF1-dependent neurite degeneration.[40] PHLDA1 promotes neuroinflammation and endoplasmic reticulum stress in neurons and is suggested a therapeutic target for neurodegeneration.[41] SCRG1 is expressed in brains of mice infected with scrapie, suggesting a role in neuroinflammation or cell death.[42] It is also suggested that SCRG1 may play a role in age- and Alzheimer’s disease-related cognitive decline.[43] (Aging astrocyte marker). TREML2 modulates neuroinflammation by regulating the NLRP3 and microglial polarization.

A comparison table of the functional categories of the periodic and aperiodic gene sets is shown in the Supplementary Table S4 and Supplementary text.

The top five genes that are most important in discriminating the clusters seem to have biological functions relevant to MCI and AD. ACAN encodes a protein critical to the structure and function of perineuronal nets (PNNs) in the brain and in Alzheimer’s disease there is a significant loss of PNNs.[44,45] Interestingly, ACAN gene polymorphism is associated with AD.[46] INPP5B is one of 50 genes whose transcriptomic profile in the brain correlates with MIND (Mediterranean-Dietary Approaches to Stop Hypertension Intervention for Neurodegenerative Delay) diet intake and calculated transcriptomic profile score was associated with cognitive decline and odds of dementia.[47] CAMKK2 is involved in post-translational phosphorylation of tau.[48] In postmortem hippocampal tissues CAMKK2 levels are significantly reduced in AD, frontotemporal dementia and parkinson’s disease, suggesting that CAMKK2 downregulation is a shared feature in several neurodegenerative diseases.[49] In addition to phosphorylation of tau, CAMKK2 has a role in calcium signaling and iron metabolism, both of which are also implicated in AD. CABIN1 is involved in synaptic plasticity, learning and memory.[50] Furthermore, in AD cohort, CABIN1 is associated with the changes in the Mini-Mental State Examination (MMSE) score.[51] SPATA13 is involved in axon growth, and synapse formation and is differentially expressed in APOE knock-in mice.[52,53] Furthermore, SPATA13 is suggested to promote dendritic spine formation and synapses in hippocampal neurons, crucial for learning and memory.[54]

#### Literature review references
